# Recovery from severe mental health problems: A systematic review of service user and informal caregiver perspectives

**DOI:** 10.1101/2021.05.13.21257172

**Authors:** Norha Vera San Juan, Petra C Gronholm, Margaret Heslin, Vanessa Lawrence, Matt Bain, Ayako Okuma, Sara Evans-Lacko

**Affiliations:** Health Service & Population Research Department, Institute of Psychiatry, Psychology and Neuroscience. King’s College London. London, United Kingdom; University of Warwick; Personal Social Services Research Unit. London School of Economics and Political Science. London, United Kingdom

**Author notes:** Authorship credits: NV participated in the conception, analysis, interpretation of data, drafting, revising, and final approval. PCG participated in analysis, interpretation of data, revising and final approval. MH participated in the conception, interpretation of data, revising and final approval. VL participated in the conception, interpretation of data, revising and final approval. MB participated in analysis, interpretation of data, revising and final approval. AO participated in analysis and final approval. SE-L participated in the conception, interpretation of data, revising and final approval.

**Keywords:** Recovery, wellbeing, severe mental disorders, service user, carer, systematic review

## Abstract

**Introduction:** The recovery approach aims to have users’ perspectives at the heart of service development and research; it is a holistic perspective that considers social needs, personal growth and inclusion. In the last decade recovery-oriented research and practice has increased greatly, however, a comprehensive model of recovery considering exclusively the perspectives of people with lived experience has not been devised.

**Aims:** This review aimed to develop a framework and contextualise service users’ and informal caregivers’ understanding of recovery from severe mental health problems.

**Methods:** A systematic search of 6 databases including key terms related to knowledge, experience and narratives AND mental health AND personal recovery. The search was supplemented with reference sourcing through grey literature, reference tracking and expert consultation. Data analysis consisted of a qualitative meta-synthesis using constant comparative methods.

**Results:** Sixty-two studies were analysed. A pattern emerged regarding the recovery paradigms that the studies used to frame their findings. Recovery domains included *Legal, political and economic recovery; Social recovery; Individual recovery;* and *Clinical recovery experience*. Service users’ definitions of recovery tended to prioritise social aspects, particularly being accepted and connecting with others, while caregivers focused instead on clinical definitions of recovery such as symptom remission. Both groups emphasised individual aspects such as becoming self-sufficient and achieving personal goals, which was strongly linked with having economic means for independence.

**Conclusions:** The recovery model provided by this review offers a template for further research in the field and a guide for policy and practice. Predominant definitions of recovery currently reflect understandings of mental health which focus on an individual perspective, while this review found an important emphasis on socio-political aspects. At the same time, only a small number of studies took place in low-income countries, focused on minoritised populations, or included caregivers’ perspectives. These are important gaps in the literature that require further attention.

**Visual abstract:** 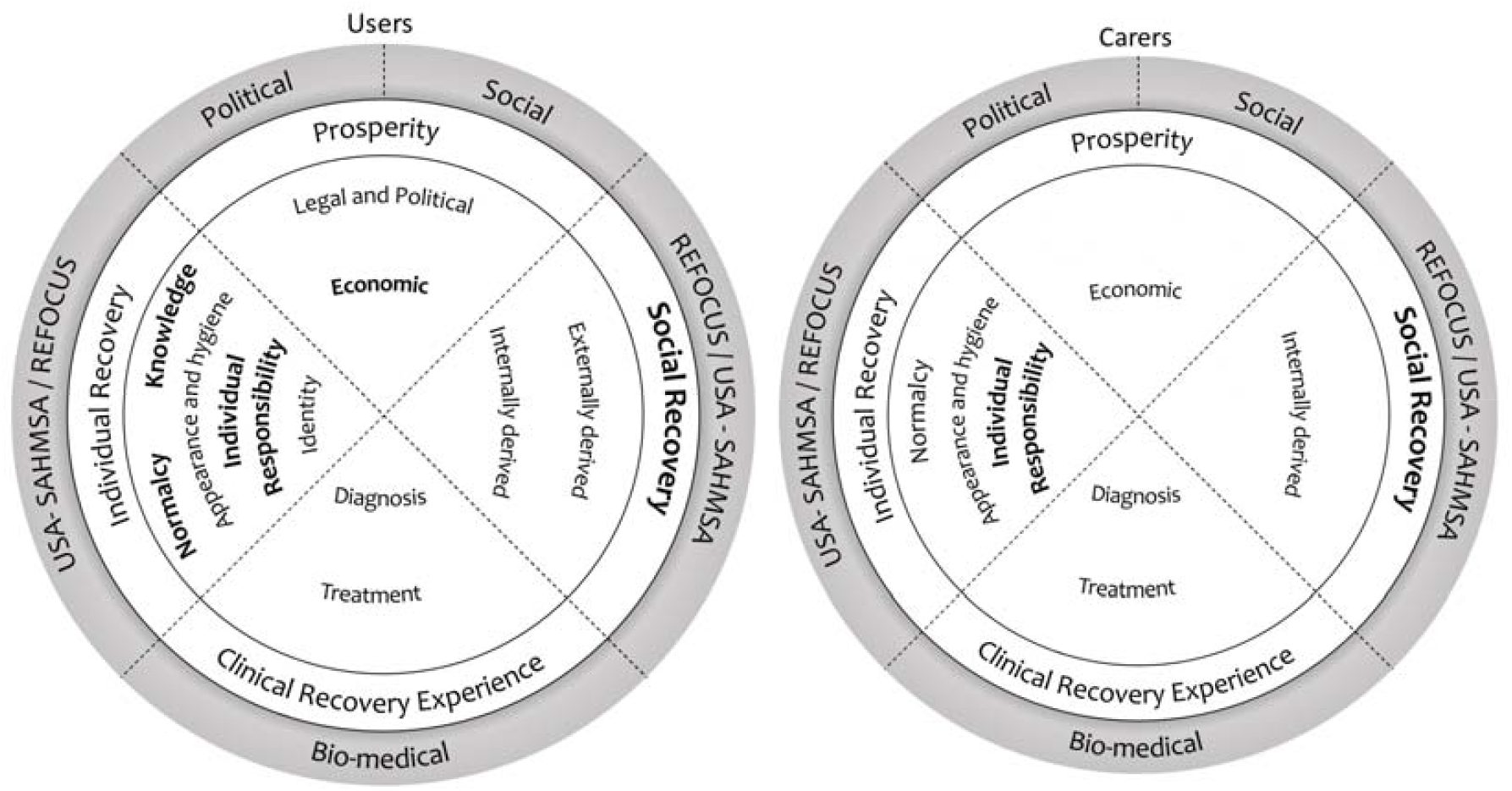

## 1.1 Introduction

The ways in which people conceptualise mental health problems vary across cultures, and therefore there are also variations in the meaning of recovery (Kleinman, 1988; Adeponle, Whitley and Kirmayer, 2012). Much of mental health practice, research and policy relies on what is known as a bio-medical understanding which speaks of mental distress in terms of diagnosis, and frames recovery in terms of clinical outcomes (John, Bentall and Roar, 2009; Pilgrim, 2009). From that perspective, recovery is focused on reduction of symptoms and functional impairment. The concept of *clinical* recovery derives from research led by mental health professionals: it involves diagnosis, and measures of symptoms and psychosocial functioning designed and rated by professionals (Slade, 2009; Piat et al., 2011). This type of recovery underpins a large number of data collection instruments that have been used in epidemiological research.

However, critics of the clinical recovery model have highlighted limitations regarding the lack of sensitivity to variability across individuals and contexts, and not including outcomes that are meaningful to service users (Crawford et al., 2011). Since the 1990’s, the focus in the field of recovery has shifted to an approach derived from literature led by mental health service users/survivors. This has been referred to as *personal* recovery, it stems from and focuses on attitudes towards life, personal growth and abilities, contribution to the community and life satisfaction (Anthony, 1993; Leamy et al., 2011). This approach aims to have users’ perspective at the heart of service development and research, and it is considered distinct from “clinical recovery” that focuses on achieving clinically-defined goals (Farkas, 2007; Piat et al., 2009; Slade, 2009; van Os et al., 2006) The personal recovery approach is an ideology that encourages a broader understanding of mental ill health experiences and how people who are feeling mentally unwell can be helped. Placing service users at the centre of decision-making in mental health has initiated a major shift in traditional philosophical views of mental health, resulting in reduced discrimination and reduced association of mental health problems with deficit and chronicity (O’Hagan, Reynolds and Smith, 2012). This definition of recovery is becoming a key concept in mental health research, policy and service development world-wide, thus progressing towards the recognition of human and civil rights of those affected by mental health problems and their carers (Bird et al., 2014).

There has, however, been criticism about personal recovery being defined in individualistic terms (Price-Robertson et al., 2017) that neglect collectivist values that are more present in some cultural groups (Chiba et al., 2010; Rose, 2014; Slade et al., 2014; Tanaka-Matsumi & Marsella, 1976). A perspective that has been lacking in conceptualisations of recovery is that of informal caregivers, whose views are not typically taken into account in recovery definitions, and thus their key role in the users’ recovery journey is not recognised. Acknowledging informal carers’ perspectives of recovery could facilitate a deeper understanding of less common paradigms which emphasise the systemic nature of recovery and take into consideration socio-economic needs and inclusion (Mezzina et al., 2006; Onken et al., 2007). Less widely cited recovery paradigms propose social and political factors to be taken into account, and add pursuing civil rights to the aims of recovery (Hopper, 2007; Pelletier et al., 2015).

In the last decade recovery-oriented research and practice has increased greatly. Recovery is now a focus world-wide and the intention to develop recovery-oriented services is typically present in official mental health service strategies (Patel et al., 2018). However, a synthesis of experts by experience’s definitions of recovery has not been devised and, therefore a comprehensive model that reflects their views is not in place. The purpose of this research is to address this gap by systematically reviewing the evidence for mental health service users’ and their informal caregivers’ understandings of recovery from mental health problems. This will allow for the development of a comprehensive model that encompasses the full range of dimensions of recovery which are relevant to experts by experience (i.e. individual and systemic recovery), while at the same time identifying key recovery paradigms and characteristics of the recovery literature to provide context for this construct.

## 1.2 Methods

This review followed the Preferred Reporting Items for Systematic Reviews and Meta-Analyses (PRISMA) statement (Moher et al., 2009). A protocol was developed a priori and registered on PROSPERO (CRD42017076450).

### 1.2.1 Search strategy and study selection

Six electronic databases (Embase, PsychINFO, Medline, ScIELO, LILACS and CINAHL) were searched in October 2020. The search strategy included key terms related to knowledge, experience and narratives AND mental health AND personal recovery. A complete search strategy is provided in Supplementary File 1. Further articles were sourced by searching for publications by authors of relevant grey literature identified in the database searches. Due to most publications identified being based in Europe and North America, a convenience sample of ten recovery experts working in seven countries across Africa, Asia and Latin America were contacted for suggestions of further literature relevant for inclusion. Additionally, the search was supplemented by reference searching through included literature, and, the five authors with most publications were contacted to enquire about potential missed studies or work in press.

Initial screening was conducted based on the titles and abstracts of the search results using the web application Rayyan (Ouzzani et al., 2016). Full texts were sourced for articles deemed relevant for inclusion and these were then screened against the full review eligibility criteria.

To establish consistency in the study selection, 300 randomly selected records at the title and abstract screening stage, and 50 records at the full text screening stage were independently reviewed by the author and a second screener, and discrepancies were resolved via discussion.

### 1.2.2 Eligibility criteria

Studies were included in this review if (1) their focus was recovery from severe mental health problems, (2) as understood by service users and informal caregivers, and (3) enquired through methodologies where participants’ perspectives were explored in an open-ended manner; studies with fixed survey responses were excluded. There were no restrictions on publication date or language.

Recovery was understood as changes towards feeling well, reaching meaningful outcomes or experiencing a positive sense of self. The term informal caregiver refers to people who provide unpaid care or support for people with mental health problems.

Articles were excluded if mental health problems were not the participants’ primary condition, or if the focus of the study was limited to a specific aspect of recovery. Studies where the primary condition was substance misuse or exposure to traumatic events were excluded due to these fields having their own extensive bodies of recovery literature which describes specific recovery paths (Davidson et al., 2005).

A full list of the inclusion/exclusion criteria is provided in Supplementary File 2.

### 1.2.3 Data extraction and risk of bias assessment

Data collected from the studies included the recovery paradigms used to frame their findings in the introduction/background section (either in terms of a paradigm explicitly stated by study authors, or a paradigm as interpreted by the review team), and the recovery themes that studies reported in the results section/discussion. When themes were not explicitly presented, results were categorised into themes. Special attention was paid to extract themes of recovery described as an outcome, rather than when presented as helping or hindering recovery. In addition, data were collected on core study details (year, setting, population and methodological characteristics, and authors’ interpretations and further discussions on the data). Missing details were requested from study authors.

Given the plurality of methodologies used in the identified studies, seven criteria for quality appraisal were adopted from different published tools (Bromley et al., 2002; *Critical Appraisal Skills Programme (CASP) Checklist*, n.d.; Dixon-Woods et al., 2004; Hong et al., 2018) with the aim of appraising transparency, description of key terms and coherence. The full risk of bias assessment checklist is provided in Supplementary File 3.

### 1.2.4 Qualitative meta-synthesis

An interpretative synthesis using constant comparison was conducted to develop a definition of core dimensions of recovery and an understanding of how they may be related (Barnett-Page & Thomas, 2009; Noblit & Hare, 1988). This method involved using reciprocal translational analysis to group the themes identified in the literature into higher order themes that best reflected their content, while keeping the theory grounded in the data and context of each study to gain a broader picture of the construct of recovery. Additionally, negative cases were kept in a log to have them present during data synthesis.

At a final stage, study characteristics were condensed into ecological sentences (i.e. “in this year, within this paradigm of recovery, in this setting, recovery meant…”) to facilitate mapping the concept of recovery (Popay et al., 2006).

## 1.3 Results

### 1.3.1 Study selection

A flow diagram of the screening and selection process, according to PRISMA guidelines, is presented in Figure 1 Preferred Reporting Items for Systematic Reviews and Meta-Analyses (PRISMA) flow diagram of the screening and selection process conducted in this systematic review.. A full list of citations and reasons for exclusion is provided in Supplementary File 4. The remaining 62 studies were included in this review.

**Figure 1.**
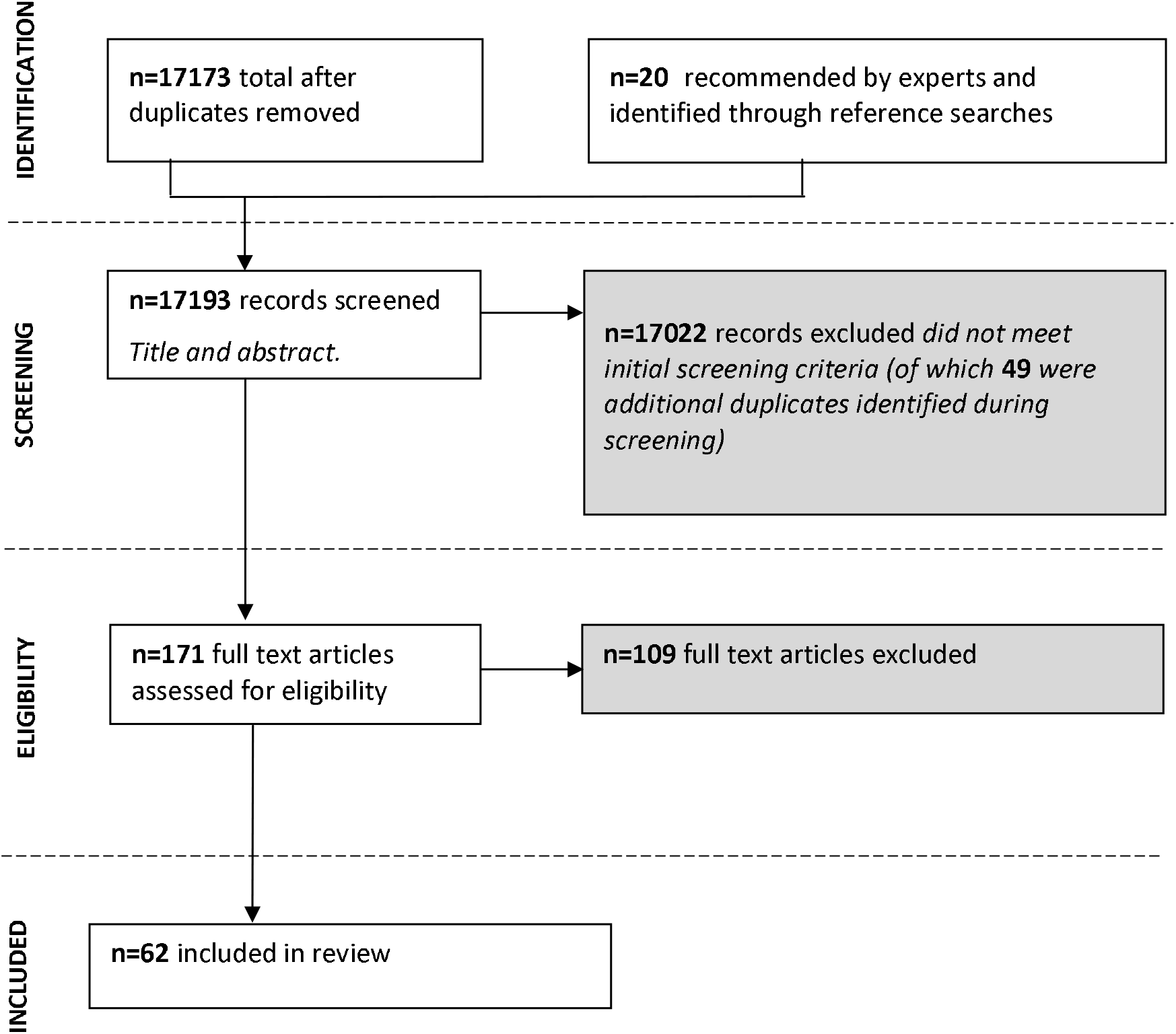
Preferred Reporting Items for Systematic Reviews and Meta-Analyses (PRISMA) flow diagram of the screening and selection process conducted in this systematic review.

### 1.3.2 Study characteristics

From the 62 papers included in this systematic review, one was published in 1967, while the rest were conducted between 1999-2020. Study settings were primarily English-speaking (n=51, 82%), high-income countries (n=58, 94%). However, six (10%) of these papers focused on a low-income sample. Recruitment was done through convenience or purposeful sampling in all studies, generally accessed through clinical contacts or announcements in recovery or service user groups.

Data were collected using in-depth interviews in 47 (76%) of the studies. Other methods included focus groups, photo-voice, ethnography field notes and narrative interviews. Thematic analysis (n=27, 44%) and grounded theory (n=11, 18%) were the most commonly used analysis methods. Two studies (3%?) applied a quantitative methodology, one followed a Delphi process for data collection and analysis (Law & Morrison, 2014), and one study used a snowball technique for data collection and Chi squared analysis (Gopal et al., 2020).

Sample sizes ranged from 1-177 participants in qualitative studies, and 180-381 in the quantitative studies. Sixty (97%) studies included a user sample, and nine (15%) included a caregiver sample. Studies typically included both male and female participants between 18-65 years of age. Twenty-five (40%) studies specified participants’ ethnicities; out of these, Nineteen were predominantly of white-European background. The remaining six studies included two in the USA and Canada which had specific interest in users of black-African descent (Armour et al., 2009; Kidd et al., 2014); one that contrasted perspectives of Euro-Canadian and Caribbean-Canadian participants (Whitley, 2016); one focused on the perspectives of women in Swaziland (Nxumalo Ngubane et al., 2019); one about Indian service users and caregivers (Gopal et al., 2020); and one focused on individuals from a Chinese community in Hong Kong (Yuen et al., 2019).

Participant information concentrated around stage of recovery and diagnosis. Authors described the stage of recovery in various ways such as length of service use or feeling well enough to participate in the study. Studies included heterogeneous transdiagnostic samples, with the exception of 17 (27%) studies that focused on psychosis/schizophrenia, 3 (5%) on depression, 3 (5%) on personality disorder, 3 (5%) on bipolar disorder, and 1 (2%) focusing on voice hearing following the single complaint approach (Bentall, 2006). Limitations were stated in relation to comorbidity with other diagnoses and relevance and usefulness of diagnostic criteria.

User employment and education were reported in 18 (29%) and 13 (21%) studies, respectively. Based on these data, users were most commonly unemployed and education levels varied from no schooling to “25 years of education”.

A pattern emerged regarding the recovery paradigms that the studies used to frame their findings. Five distinct categories were identified: USA consumer/survivor recovery movement (including Substance Abuse and Mental Health Services Administration – SAMHSA-model) (n=19, 30%); REFOCUS-CHIME model of recovery (n=12, 19%); Social recovery (n=8, 13%); Political recovery (n=3, 5%), and Bio-medical recovery (n=3, 5%). Recovery paradigms concurred in acknowledging the potential to feel better after experiencing mental health problems, however, they differed in their position regarding four aspects of recovery: (1) The extent to which they focused on internal conditions such as individual’s attitudes, versus external conditions such as policies and social circumstances; (2) the importance placed on diagnosis; (3) the literature by which they were influenced, and thus (4) the recovery goals they proposed to focus on. A brief description of each paradigm is provided in Table 3. The main characteristics of the included studies are listed in Supplementary material 5.

### 1.3.3 Risk of bias

All studies met 50% or more of the quality criteria assessed, and 31 studies (50%) fulfilled all 7 criteria. Additionally, a substantial number of studies included user participation or mindful interviewer selection (n=29, 47%) to enhance rigour.

## 1.4 Recovery themes

This list of themes is the result of the synthesis of the empirical data extracted from the results section of the studies included in this review. Table 2 illustrates the four core parent themes present in these data: (1) Prosperity; (2) Social Recovery; (3) clinical recovery experience; (4) individual recovery. All themes were present to a greater or lesser extent in users’ definitions of recovery; the cases where themes were also part of caregivers’ understanding of recovery are highlighted where applicable. These themes are elaborated upon below, with selected quotes from the included studies illustrating the key characteristics of the parent themes and subthemes within these. Figure 2 provides a visual representation of how the findings in this review are related. Theme one was aligned with the social and political recovery paradigms, theme three with the bio-medical recovery paradigm, and themes two and four overlapped with the definition of recovery of the REFOCUS-CHIME, SAHMSA and USA consumer/survivor movement. At the same time, social and political aspects of recovery were more common among user samples, while clinical recovery goals were more prevalent among carer samples.

**Table 1.**
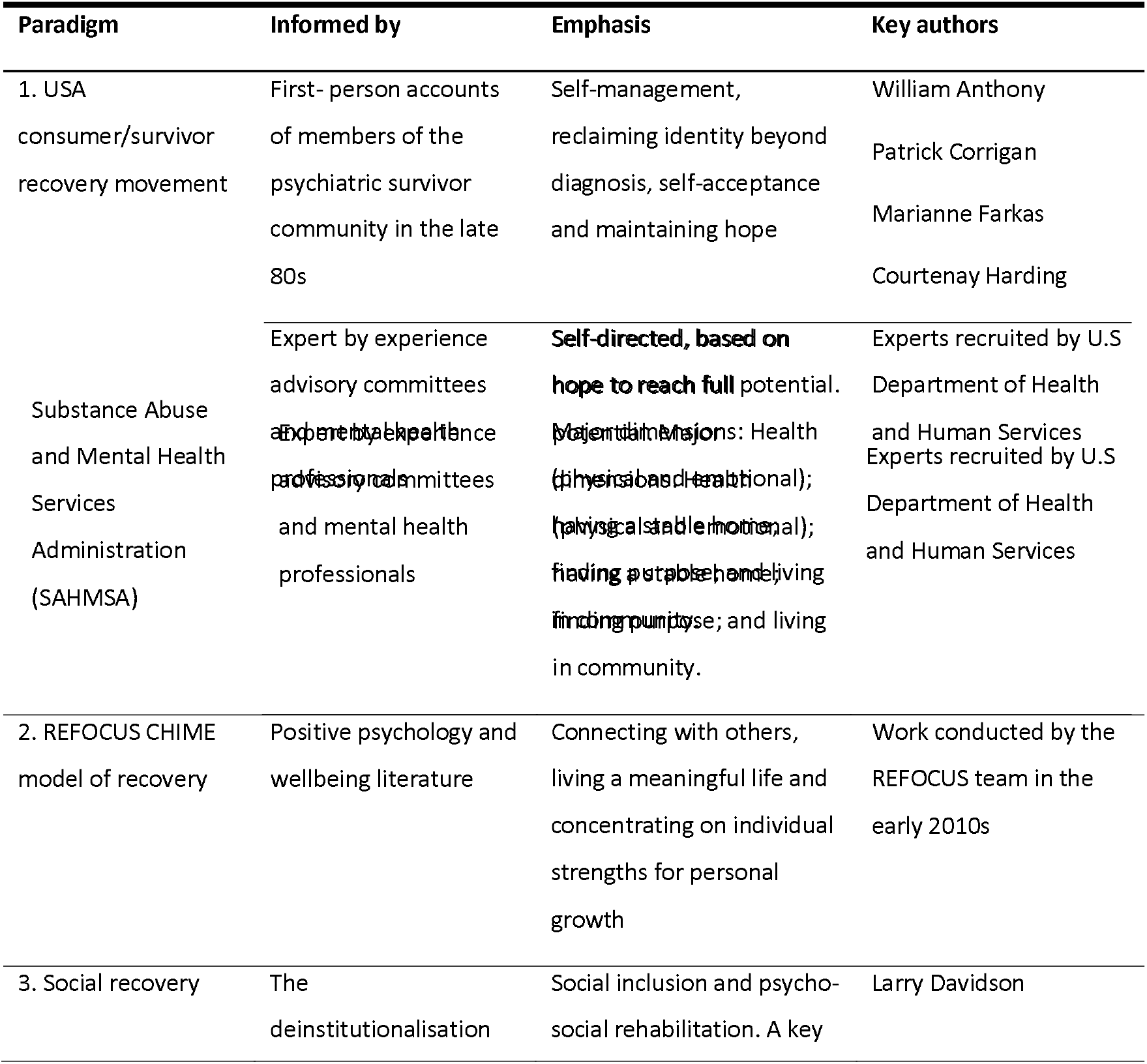
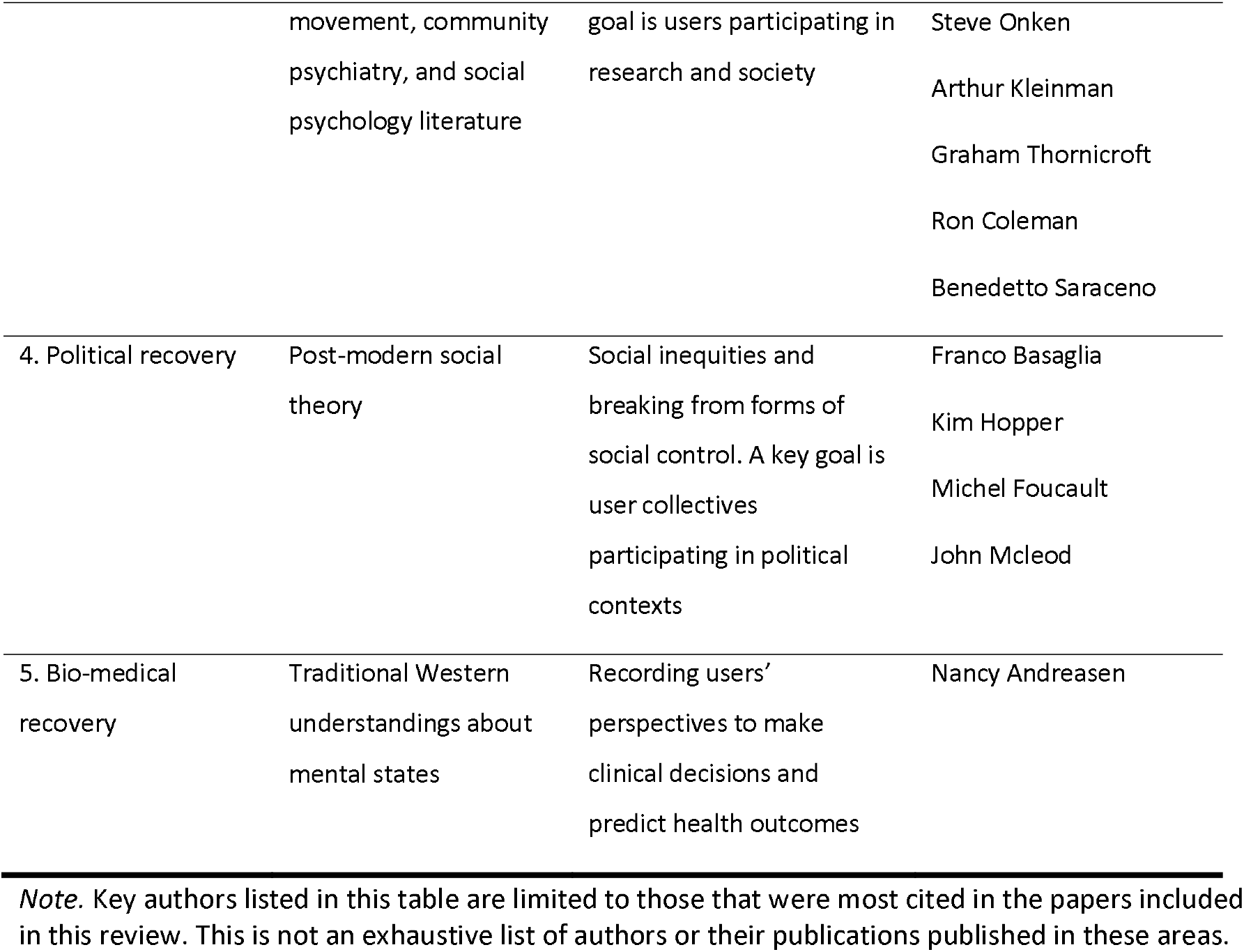
Description of recovery paradigms identified in the literature.

**Table 2.** Parent themes identified in the data, the subthemes that fall within these and the number of user/carer studies which included them.

| Parent Theme | Subthemes | N (U=53/ C=9)* | Description |
| --- | --- | --- | --- |
| Prosperity | • Legal and political recovery | 7 / 0 | Linked to empowerment; covering basic economic needs and co-construction of recovery |
|  | • Economic recovery | 21 / 2 |  |
| Social Recovery | --- | 41 / 4 | Returning to a basic form of social awareness; being a part of society, functioning well within groups, treated as an equal |
| Individual recovery | • Normalcy | 21 / 2 | Being 'normal'; completing everyday activities and/or focusing on achieving personal goals; fulfilling roles and responsibilities; gaining relevant knowledge about mental health or enrolling in formal education. |
|  | • Temporal understandings and identity | 21/ 0 |  |
|  | • Recovery and knowledge |  |  |
|  | • Recovery as an individual responsibility | 49 / 5 |  |
|  | • Appearance and hygiene | 4 / 1 |  |
|  | • Recovery as a positive frame of mind | 19 / 2 |  |
| Clinical recovery experience | --- | 17 / 5 | Considerations about diagnosis and treatment |

\*This column indicates the number of User/ Carer articles that included each theme. Articles with a User sample total N=60; articles with a carer sample total N=9.

**Figure 2.**
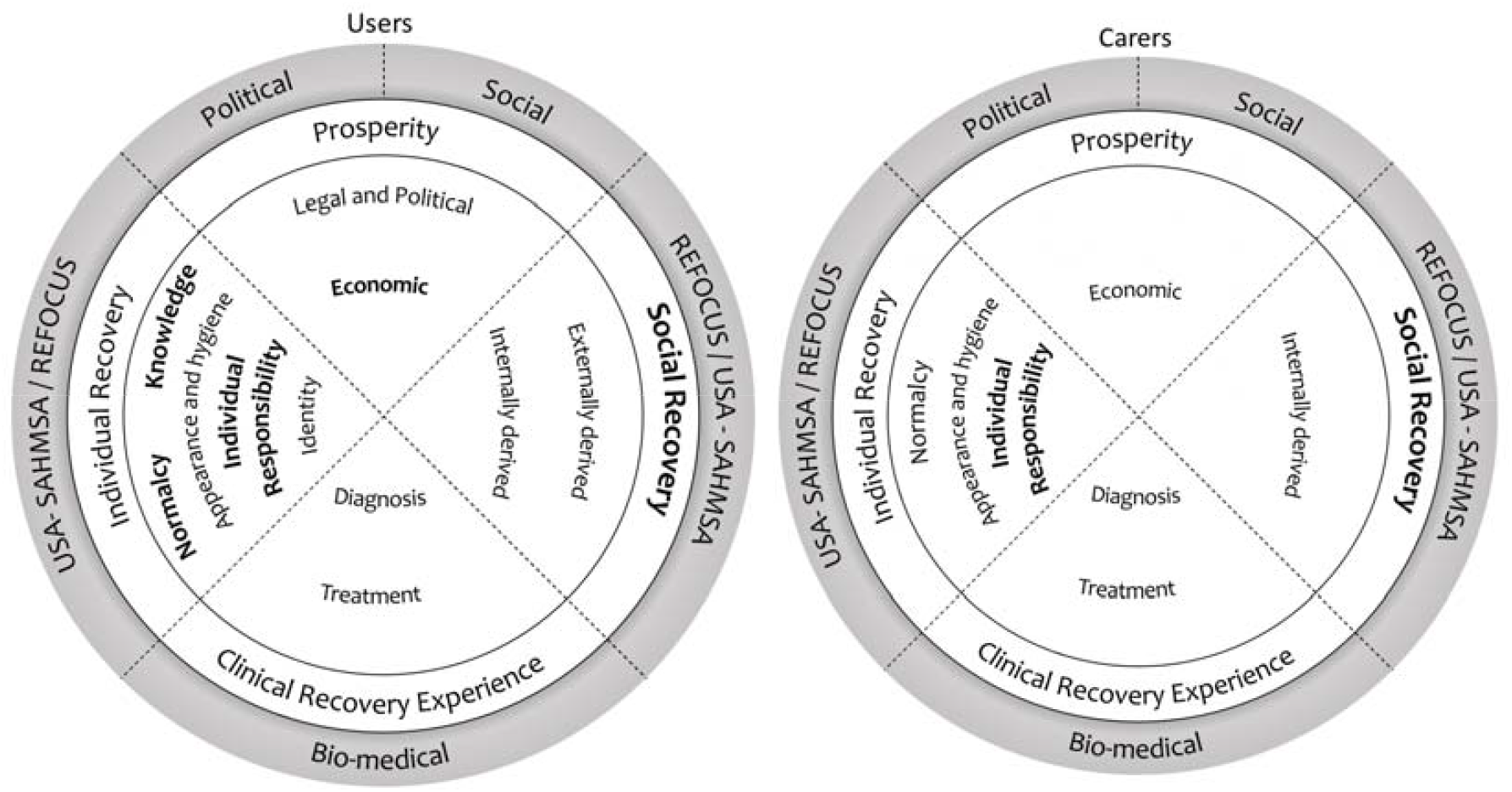
Meta-synthesis map. *Note*. Visual representation of how the recovery paradigms and themes identified in this systematic review are related and their predominance in user/carer samples. The circles on left and right represent recovery as understood by users and carers respectively. The outer circle presents recovery paradigms, while the inner circles refer to the themes and subthemes. The most prevalent themes are highlighted in bold letters.

### 1.4.1 Theme 1: Prosperity

Framing recovery as a social construct was highly present in the literature. Examples of this can be found in Basso et al. (2016) “*recovery has to be understood also as a social process, where people face, along with the disease, other tests such as the need for tangible resources, jobs, availability of housing, financial independence, and efficient services” or* *Kidd et al*. (2014), who studied recovery from the perspective of racialised women in Canada and remarked on the lack of discussion around symptoms and treatment in participant’s recovery narratives: “*their challenges were very much framed as social rather than psychiatric”*.

This recovery theme was especially common in literature linked to the user/survivor movement or advocating for collective action against human rights violations in mental health treatment.

Subthemes that fell under this theme were: “Legal and political recovery” and “Economic recovery”.

#### 1.4.1.1 Legal and political recovery

Empowerment was one of the central aspects underlying this theme; recovery goals were related to rebelling against socially imposed rules or practices which users considered to stand in the way of their wellbeing and advocating for fairer legislation. This idea was especially prominent in the literature analysing women’s understanding of recovery, where these thoughts were discussed under the terms “*breaking away from limited woman roles”* (*Kidd et al*., *2014)* *and “doing and being beyond gendered responsibilities”* (Fullagar & O’Brien, 2014). Fullagar and O’Brien (2014) concluded *“Practitioners and advocates in women’s health movements have historically recognised that personal recovery is political.”*. At the same time, Armour, Bradshaw and Roseborough (2009), pointed out that black and minority ethnic (BME) groups experienced oppression both because of their mental health problems and because of their race, which would involve two different approaches when fostering empowerment.

#### 1.4.1.2 Economic recovery

A key recovery goal from both a user and caregiver perspective was reaching economic stability. Recovery was understood as having sufficient resources available to have an acceptable quality of life and live independently from family. Participants in Borg & Davidson’s (2008) study in Norway, included shopping and paying bills as part of their notion of achieving “normality” (see *normalcy* subtheme). Similarly, service users and carers in Italy considered recovery involved actions to reduce external barriers that impeded independent living, such as lack of jobs in the open market and lack of accessible living solutions which prolonged cohabitation with the family (Basso et al., 2016). The need for financial support and/or access to employment to mitigate adverse material circumstances was highlighted particularly in studies with participants from ethnic minorities or hard to engage populations (Armour et al., 2009; Milbourn et al., 2014; Nxumalo Ngubane et al., 2019).

### 1.4.2 Theme 2: Social recovery

Two interrelated types of social recovery were identified. One was an externally derived social recovery which required approval and acceptance from the group. In this sense, recovery meant being trusted, being assigned responsibilities and being treated as an equal. Cárcamo Guzmán et al., (2019) wrote about the meaning of recovery to service users in Chile, “*it is understood as the legitimacy of the user as a person, this implies the respect for their experiences, points of view and needs*”. The other type of social recovery was derived from personal initiative and consisted of: socialising and establishing meaningful relationships, being a productive member of the community, and fulfilling family roles. Participants in Hancock, Smith-Merry, Jessup, Wayland, & Kokany’s (2018) study spoke about learning to navigate complex relationships, avoiding unhelpful interactions and managing the impact of their mental health problems on others.

Nxumalo Ngubane, McAndrew, & Collier, (2019) presented being accepted and able to contribute to their family and community as an important part of recovery for Swazi women diagnosed with schizophrenia. The socially constructed nature of recovery was emphasised repeatedly, with social discrimination and experiences of stigma being perceived as the opposite of recovery in many of the studies (Cárcamo Guzmán et al., 2019; Gopal et al., 2020; Lee et al., 2020; Nowak et al., 2017; Nxumalo Ngubane et al., 2019; Shepherd et al., 2017; Tofthagen et al., 2017). The definition and achievement of recovery was thought to be co-constructed in society and developed by engaging in honest and genuine mutuality (Kverme et al., 2019). In this sense, others offering help or feedback, and users being willing to accept it, were equally important recovery goals, as pointed out by Moltu et al. (2017) in Norway saying *“In our analyses, we were struck by how important others were in noticing improvement and positive change, in a way that the suffering person could embody”*.

An important part of externally derived social recovery was being allowed to take risks, this is to be considered to have adequate judgement in everyday life and legal capacity to consent in formal contexts. As written by Pitt et al. (2007) *“ultimately recovery requires active participation in life. This involves taking risks and suffering setbacks.”*. Fullagar and O’Brien (2014) described how an environment that allowed for free decision-making provided users with the opportunity to experience “*dignity of risk*” and realise their capabilities.

Some studies described a spiritual form of connection with a *“higher power”* or *“God”* as important for recovery (Armour et al., 2009; Nxumalo Ngubane et al., 2019; Ochocka et al., 2005). Allusion to spiritual or religious recovery concepts was present across the literature in the different populations and settings.

People with mental health problems which affect social interaction, such as people with a diagnosis of personality or bipolar disorder, were thought to face a greater challenge to achieve social recovery. This was both related to personally derived social recovery, as described by Katsakou *et al*. (2012)*”improving relationships for this group might also be more complex than solely addressing social isolation* [discrimination], *which is commonly discussed in recovery literature”*, and externally derived social recovery, Kverme et al. (2019) *“The experience of becoming safer as a human among other humans constituted a core meaning of recovery”*.

Within this theme, caregivers’ definitions of recovery concentrated mainly around users being attentive to others’ needs and able to establish positive connections. As mentioned by Tweedell *et al*. (2004) “*Families described changes in amount and content of interaction, noted their relative being helpful in the home, showing consideration for a parent, remembering a family member’s birthday*.”, and by Yuen, Tse, Murray, & Davidson (2019) “*She* [carer’s daughter] *can integrate into society through such things as going to church, having a job, returning to a normal life, going out’*”.

### 1.4.3 Theme 3: Individual recovery

The third parent theme focused on individual goals, needs and responsibilities. As expressed by Young and Ensing (1999) *“Contrary to the common belief that mental illness involves a purely degenerative condition, it appears that many people discover new potentials and new self-growth at various points throughout their recovery.”*

This theme of individual recovery encompassed six subthemes: “Normalcy”; “Temporal understandings of recovery and identity”; “Recovery and knowledge”; “Recovery as an individual responsibility”, “Appearance and hygiene” and “Recovery as a positive frame of mind”.

#### 1.4.3.1 Normalcy

Related to social recovery was the idea of not feeling different from most people and achieving the goals that are considered the norm by your social group. Borg & Davidson (2008) found “*being normal*” to be one of the major themes in recovery: “*What seems most crucial to ‘‘being normal’’ is spending time in ordinary environments with ordinary people*.”. Katsakou et al. (2012) identified a link between employment and feeling normal, as expressed in one of their participant’s quotes: “*I still haven’t managed to get back to work and I can’t see friends, I’ve been cut off because I’ve stopped working*”.

A line of the recovery literature focused on understanding recovery through ordinary everyday activities. In this sense, the main recovery goal consisted of completing routine tasks and participating in common leisure activities. Milbourn, McNamara and Buchanan (2014) noted that in order to appreciate participants’ understandings of recovery, the list of everyday routines needs to be broadened to include personally meaningful activities which may be considered negative by others, such as *“recreational drugs and paying for sex”*.

McCabe, Whittington, Cramond, & Perkins (2018) pointed out in forensic mental health services *“everyday activities such as walking and discussing books were talked about in the language of therapies administered by services. The ‘reader group’ and the ‘walking group’ were all discussed in terms of therapeutic interventions rather than fulfilling hobbies that people adopt in everyday life”*.

#### 1.4.3.2 Temporal understandings of recovery and identity

There were two contrasting views regarding the temporal focus of the recovery journey: one described recovery as the ability to focus on future goals, having hope and seeing “*the light at the end of the tunnel”* (Vander Kooij, 2009), while the other described it as the ability to live in the present and complete daily routines (related to the “Normalcy” subtheme). This contrasting view of recovery was also present in different identity goals, with some service users striving to develop a ‘new self’ by learning from their experience (Leavey, 2005; Ridge & Ziebland, 2006), and others wanting to return to the roles and occupation or everyday activities from before experiencing mental health problems (Hipolito et al., 2011; Tweedell et al., 2004). Recovery was not a single state of being but a complex mix of the past, the here and now and, the future (McCabe et al., 2018).

This distinction was discussed by de Jager *et al*. (2016), who found participants wishing to reflect on and integrate the disorder experience into a new identity, while others wished to leave the experience behind and focus on symptom management. Participants in both groups were described as currently not having symptoms, good quality of life and no psychological distress, for this reason the authors advocated for the latter approach to recovery to not be pathologized. Instead, they promoted a broader understanding of recovery that does not require active engagement or reflecting on the mental health problems experienced.

The idea of returning to a former identity was a prominent topic throughout the literature, however, it was particularly highlighted in the definition of recovery of older adults presented by Daley et al. (2013)”*The single core category identified from the analysis was ‘Continuing to be me.’ This related to the permanent and established sense of identity which service user participants held […]*.”.

#### 1.4.3.3 Recovery and knowledge

An important recovery goal was gaining new knowledge. This included knowledge about yourself (personal growth), knowledge about mental health, and knowledge gained through formal education. The latter was highlighted as particularly important in Simonds *et al*. (2014) study about adolescent service users.

Service users in Shepherd et al.’s (2017) study underlined the role of understanding early lived experience as informing sense of self “*Most participants framed their understanding of their experiences within a description of their early life within their family, particularly their sense of belonging and the interpretations of their behaviour made by key family members*”. Self-discovery was also a significant part of recovery for young people in McCauley, McKenna, Keeney, & McLaughlin’s (2017) study, pointing to the limited life experience before mental health problems creating an additional vulnerability.

Knowing more about mental health was approached both as part of embracing a given diagnosis (e.g. the goal “*coming to know your illness”* (Yarborough et al., 2016) and discarding it (e.g. “*developing a critique of mental health services”* (Pitt et al., 2007). These considerations about diagnosis are explored further later under the theme “Clinical recovery experience”. In both cases the final aim was to develop strategies to feel better, building higher self-esteem and self-awareness. As described by Engqvist and Nilsson (2014) “*Recovery usually occurs when people with mental disabilities discover or rediscover their strengths and the opportunities to pursue personal goals and a sense of self that allows them to grow, despite any residual symptoms and difficulties*.”.

#### 1.4.3.4 Recovery as an individual responsibility

Being self-sufficient and having control over one’s mental health problems and their consequences were highly prevalent recovery goals. Recovery within this theme is described as an internal fight, coming to the realisation that *“It needs to be me”* (Hancock et al., 2018). In most of the literature, recovery was presented as a personal choice to actively cope with mental health problems. An important aspect of reaching autonomy was no longer being reliant on mental health services. As stated in Todd, Jones and Lobban (2012), participants did not consider mental health services to promote self-management and this was seen as going against their recovery. Participants wished to assert their position as experts by experience and those who did not engage with services were seen as “winners”: *“taking responsibility is at the heart of the recovery process as people are empowered to make their own choices and focus on their own outcomes.”*.

This understanding of recovery is summarised by a participant in the study conducted by Mizock, Russinova and Shani (2014)”*“It reminds me of an author who said she’s never avoided challenges but put her “sails full tilt into the wind.” There’s a certain bravery in facing obstacles head-on. With my mental health challenges, I’ve learned to put my sails full tilt to the wind and move towards my goals*.”.

This conceptualisation of recovery as an individual responsibility was strongly linked to empowerment, which in turn was linked to having economic means for independence (Basso et al., 2016; Brijnath, 2015; Santos et al., 2018). A person with mental health problems reaching independence was a particularly important recovery goal for caregivers, this included financial autonomy and independent living that reduced the reliance on caregiver/family support (as mentioned in the subtheme “Economic recovery”) and reaching emotional stability. An example of this are the findings from the study by Tweedell *et al*. (2004): “*They longed for their relative to be able to take care of themselves, live independently, or have improved judgment and concentration, or to work and become functional and self-sufficient*”.

A distinctive understanding of recovery was presented by Mezey et al. (2010) who studied the views of forensic psychiatric patients (offenders with mental health problems). For the most part participants chose to rely on medication and medical guidance, rather than their own judgment and active participation: *“Their lack of control was in most cases, simply stated as an incontrovertible fact.”*

#### 1.4.3.5 Appearance and hygiene

Some studies described improving appearance and keeping up good hygiene as part of personal recovery; the focus of this goal was adding to a personal sense of worth, rather than complying with social rules. Davis (1967) who conducted an ethnography in a women’s psychiatric ward noted *“Wearing their own clothing again adds to their appearance of well-being. […] this makes it all the more difficult for them to see themselves as “sick persons.”*. A participant in the study by Santos et al. (2018) expressed *““[I want to] maintain…good hygiene…, fitness, exercise, nutrition…*””.

#### 1.4.3.6 Recovery as having a positive frame of mind

A representative description of this understanding of recovery can be found in Kartalova-O’Doherty and Tedstone Doherty (2010a) *“Personal definitions of recovery fell into two broad areas: getting rid of negative feelings, such as anxiety, depression, or panic attacks; and acquiring positive feelings and actions, such as peace of mind […]”*.

Accounts of recovery found in the literature that fall within this category include “*being positive”* (Gillard et al., 2015), “*being happy and successful”* (Kartalova-O’Doherty & Tedstone Doherty, 2010; Simonds et al., 2014), *“finding hope and purpose”* (Hancock et al., 2018), or “*having a meaningful and satisfying life”* (Yarborough et al., 2016), without a deeper description about what this meant. Recovery was described as general feelings and attitudes that were considered positive or the opposite of being unwell, dissatisfied or unsuccessful.

Another important aspect within this theme was the idea of recovery as having peace of mind (Kartalova-O’Doherty & Tedstone Doherty, 2010; Vander Kooij, 2009; Young & Ensing, 1999). This was described as feeling at ease, enjoying leisure moments or not experiencing constant anxiety and fear.

### 1.4.4 Theme 4: Clinical recovery experience

This theme includes topics traditionally related to clinical understandings of recovery such as diagnosis, medication and symptom-related concerns. Examples when this was present in the literature were references to recovery goals such as *“chemical balance”* (Ridge & Ziebland, 2006), *“adherence to treatment”* (Mizuno et al., 2015), or *“reducing clinical symptoms”* (Cárcamo Guzmán et al., 2019; Nowak et al., 2017; Simonds et al., 2014; Wood et al., 2010). Brijnath (2015) challenged traditional personal recovery literature writing: *“Participants’ emphasis on being ‘cured’, achieving an endpoint in their depression and discontinuing medicines runs counter to the recovery discourse that emphasises that one can be ill and still live a meaningful, contributory life”*. In the same line, Piat *et al*. (2009) remarked that *“The prominence of the illness perspective of recovery among consumers was unexpected. Many looked for recovery outside of themselves: in a cure, or in dreams of disappearing symptoms.”*.

For service users in some studies, recovery meant being discharged. This in turn had implications for recovery milestones being prioritised by participants, as described by McCabe et al. (2018) *“service users identified their relationships with staff as of greater importance than those with other service-users […] attaining discharge was a more immediate and pressing goal and staff were seen as holding the key to discharge […] In order to be deemed to be recovering service users were keen to demonstrate an acceptance of the bio-medical model regardless of whether this actually fitted with their view of the world.”*

With regard to diagnosis, there were two opposing views: recovery as embracing the label and recovery as dropping the label. For the first, Ridge and Ziebland (2006) used the term ‘coming out of the closet’, since accepting the given diagnosis was understood as way to achieve authentic living without trying to pass as ‘normal’. Assimilating the diagnosis as part of one’s identity also meant giving central importance to complying with treatment and medication. Brijnath (2015) found that Indian participants found meaning in life through religion, while *“For Anglo participants, meaning in life was derived from the illness experience itself. Participants talked about the importance of a diagnostic label in validating how they felt, discovering their inner strength and learning to live with depression”*.

In contrast, recovery as a rejection of the given diagnosis usually implied disengaging with services. This view was especially prevalent in literature from the user/survivor or feminist movements, and it was linked to poor practices of mental health services. Examples can be found in Adame and Knudson (2007) *“Another traditional construction from the survivors’ narratives was “recovery from the mental health system” […] all four participants felt that recovering from psychiatric interventions (e.g*., *ECT, drugs, solitary confinement) was one of, if not the biggest, challenge in their entire healing process”* and in Nxumalo Ngubane et al. (2019), where participants believed health professionals, traditional healers and religious leaders had used labelling as a form of coercion to support their own ideas of recovery.

At the same time, some studies found both views represented in their sample, such as Shepherd, Sanders and Shaw (2017) who studied recovery in people diagnosed with personality disorder and concluded that most found it useful and *“For a minority of participants however the diagnosis of personality disorder was seen as unhelpful - representing a direct comment on them as a person, or as a representation of their previous behaviour, not a ‘mental illness’ per se”*.

Clinical understandings of recovery were particularly common among carers (it was the predominant theme in five out of the nine papers that presented caregivers views) and it was normally presented as part of the guidance they received from their psychiatrist. To this respect Jacob, Munro and Taylor (2015) wrote “*Even though carers are the closest people that many consumers have in their life, carers had major divergence in their views on mental health recovery. Contrasting to consumers and nurses, none of the carers described regaining one’s sense of self as an important aspect to mental health recovery. The carers’ views on mental health recovery closely related to the traditional views of remission of symptom*”. Also, the same study reported that of importance was that this understanding of recovery led caregivers to think recovery was impossible as they understood these goals (e.g. symptom remission, retuning to pre-illness status) as unattainable: *“‘I don’t understand what you mean by recovery from mental illness, there isn’t one we went to the psychiatrist the other day and she said [that] the illness will never go’”*.

## 1.5 Discussion

This review aimed to define the various ways in which service users and carers conceptualised recovery and to provide context for how this construct is represented in the existing literature. Data from sixty-two studies originating mainly from high-income countries were synthesised and analysed. The most prominent themes in users’ definitions of recovery were Social Recovery and Individual Recovery. Within these themes, users’ understanding of recovery revolved especially around connecting with others, and recovery as an individual responsibility to reach control over mental health problems. In the case of informal carers, the most common themes when defining user recovery were Recovery as an Individual Responsibility, particularly reaching autonomy/being self-sufficient, and Clinical Recovery Experience, mainly symptom remission. Marshall *et al*. (2013) also found informal carers had pessimistic views about the potential for recovery and emphasised clinical aspects of recovery. As a possible solution they pointed to recovery training which has been found to be effective among staff (Salgado et al., 2010) and could perhaps be mirrored in carer populations.

Service users’ perspectives overall resonated with the more established models and definitions of recovery mentioned in the introduction (Anthony, 1993; Leamy et al., 2011) and identified as paradigms 1 and 2 in Table 1. These definitions of recovery are present in the themes “Individual recovery” and “Social recovery” (derived from personal initiative) proposed in this review, which focuses on personal growth, autonomy and individual initiatives. This is consistent with a review of user autobiographical accounts provided by Drake and Whitley (2014), who concluded that recovery was “*a growing sense of agency and autonomy, as well as greater participation in normative activities, such as employment, education, and community life*”, or the study conducted by Boumans *et al*. (2017) who wrote “*For our participants, successful living is fundamentally connected to “not being dependent on mental health care”*“.

However, along with providing further evidence in support of previously defined models and definitions of recovery, this review identified additional dimensions, namely social (externally derived), political and economic aspects of recovery and factors related to social reciprocity and acceptance. These understandings of recovery were consistent with less prominent recovery paradigms (3 and 4 in Table 2). This is consistent with the findings of the systematic review conducted by Llewellyn-Beardsley et al. (2019) to synthesise typologies of user recovery narratives. The authors found that recovery narratives incorporated social, political and human rights aspects to a greater extent than illness narratives. Petros *et al*. (2016) suggested an adaptation of the REFOCUS-CHIME model of recovery (paradigm 2 in Table 2) to underline the bi-directional nature of recovery. To this respect they wrote “*perceived reciprocity within […] relationships is correlated with higher levels of satisfaction in support and higher levels of personal confidence, self-esteem, and perceived recovery*”. The integral role in personal recovery of family and community has been especially mentioned in literature referring to cultures that focus more on group goals than on self-responsibility (Adeponle et al., 2012; Chiba et al., 2010). An example of this is Mak, Chan, & Yau (2018) including the domains “family involvement” and “social ties and integration” as part of their scale to measure personal recovery in Chinese culture.

Furthermore, an emphasis on availability of basic needs as exemplified in the theme “Economic recovery” was also found to be a key concern for users in the review conducted by MacÍas (2011) and the Australian National Survey of Psychotic Illness (Morgan et al., 2012). The importance of factors related to social justice which fall under the theme “Prosperity” is widely supported by research on social determinants of health (Morrow & Weisser, 2012; Verhaegh et al., 2011; Wickham et al., 2014).

There has been substantive criticism about the field of recovery being excessively focused on the individual (Price-Robertson et al., 2017; Rose, 2014; Spandler et al., 2015); researchers have raised awareness on the risk of glossing over important social challenges and the stressful social conditions that can be generated by high expectations of self-control in adverse contexts (Myers, 2009, 2010). Yates, Holmes, & Priest (2012) addressed this gap in recovery literature by studying in detail the social and environmental conditions in which recovery takes place, concluding recovery should be understood as an interaction of ecological processes such as the co-occurrence of personal growth and self-determination in contexts of social structures that restrict personal agency.

Thus, addressing social, political, and economic disparities and opportunities for participation in the community should also be recognised as a key dimension of recovery. This discussion is especially relevant for the development of the recovery approach in low- and middle-income countries (LMICs) that are affected to a greater extent by social inequality, violence, or other social stressors (Annan et al., 2013; Rauchfuss & Schmolze, 2008). Despite identifying a limited amount of research from LMICs that focused on recovery, the key role of economic sufficiency, housing, and respect of basic human rights in mental health are highly present in literature relating to both LMICs and BME groups (Blignault et al., 2009; Câmara & Ornellas Pereira, 2011; Sanders-Phillips, 1996). It has been the focus of recent calls for a paradigm change in the field of global mental health (Cosgrove, Mills, et al., 2020; Cosgrove, Morrill, et al., 2020; Mezzina et al., 2019), particularly in the context of the COVID-19 pandemic (Kola et al., 2021).

Another aspect of dominant definitions of recovery that is contested in the findings of this review is the high prevalence of clinical notions of recovery. The theme “Clinical Recovery Experience” highlighted how topics traditionally considered to fall under *clinical* rather than *personal* recovery are important aspects of users’ everyday lives and notion of recovery. Despite an attempt in the recovery-oriented discourse to diverge from “clinical” language and make a clear-cut distinction between “clinical” and “personal” recovery, our findings showed the presence of clinical concepts in users and carers understandings of recovery. However, there was a distinctive social meaning behind the clinical language of users and carers. there is also a need to study the meaning of clinical language when used by lay stakeholders in order to further understand the role that it plays in their individual and social recovery. This disparity between a social and a clinical understanding of clinical language has great importance for the development of meaningful mental health evaluation tools and clinician-user communication. This would affect decisions such as that made by Mak, Chan, & Yau (2016) of removing items related to symptom management and medication from a personal recovery measure.

Regarding diagnosis, the criticism about the lack of validity and practical use of diagnostic categories expressed in the background literature of the included studies contrasted greatly with the notable adherence to the diagnosis identity on the part of users and caregivers. Some authors have highlighted the social role of diagnostic labels, such as Cruwys and Gunaseelan (2016) who found that people diagnosed with depression tended to identify more with their diagnosis when they faced stigma, using the identification with a group as a buffer against discrimination. Tekin (2011) pointed to risks of diagnosis being a ‘double-edged sword’ that on one hand may facilitate self-understanding and communication, while on the other hand may lead users to make sense of situations focusing only on unrealistic dichotomous outcomes. At the same time, some researchers have suggested there may be an excessive representativeness of user narratives which align with medical views due to user samples consisting for the most part of responsive persons who are in a disempowered position (Castillo et al., 2013; Gillard et al., 2015; Rector, 2009).

### 1.5.1 Implications

Service user and carer accounts reviewed in this study show experiences of severe mental health problems are multifaceted and require an ecological/holistic approach. In light of these results, efforts in mental health policy and service development should address users’ social and legal disadvantages and economic distress. Articulating a civil rights or social work perspective on recovery from mental health problems would help to meet the recovery goals presented as most important to service users.

With respect to practice, worrying levels of stigma and discrimination in psychiatric practice were identified in users’ testimonies and reflected in caregivers’ notion of recovery. These are direct barriers to recovery and therefore there is a pressing need to consider the negative effects that narrow medicalised attitudes have on people’s lives. At the same time, the legal or social barriers that prevent psychiatrists from promoting user freedom and participation should be addressed (Gambino et al., 2016; Price-Robertson et al., 2017; Sartorius, 1998). Clinical and personal recovery are intrinsically related and can complement each other; optimal provision of services can be achieved by combining the strength of professional’s knowledge and epidemiological research, with stakeholder’s experience and feedback about their needs (Van Eck et al., 2018).

The particular understandings of recovery identified in this review would benefit from specific therapeutic techniques. Service users who underlined the importance of bi-directional communication for recovery may adhere better to treatments of a dialogical nature (Moltu et al., 2017), while users less interested in active engagement and meaning-making, such as those searching to achieve normalcy through completing everyday routines, could find more use in mindfulness-oriented techniques (Barnhofer & Crane, 2009; Siegel, 2010). In the same way, service users expressing concerns relating to discrimination, legal and economic circumstances should be referred to appropriate help which focuses on facilitating access to adequate housing, employment, education and money management, to ultimately be empowered to address their needs (Elbogen et al., 2011). Examples of this are initiatives such as the Bapu Trust for Research on Mind and Discourse, in India (Davar & Dhanda), and advice services set by government in the United Kingdom such as the Money Advice Service. Altogether, identifying users’ personal recovery goals and mapping them onto the framework proposed in this review would in turn facilitate the development of person-centred individualised care.

There is a need for research about recovery across different cultures. Predominant definitions of recovery currently reflect Western understandings of mental health which focus on an individual perspective, without adequately addressing important socio-political aspects. Recovery-oriented research and practice should take an additional step beyond focusing on what occurs in clinical settings and empower communities for the promotion of human rights, thus shifting from questions around why addressing socio-political recovery to how we can address user’s holistic wellbeing.

At the same time, only a small number of studies included caregivers’ perspectives. Findings from these studies suggest the recovery approach has not yet permeated this group’s view, and further attention to informal carers in research would be a step towards recognising their potential to contribute to mental health care and users’ wellbeing. Users and caregivers should be included as partners in the development of knowledge and services to ensure their personal needs and external challenges are accounted for and met.

Lastly, research into recovery identified in this review demonstrated important characteristics that helped to mitigate bias. Studies benefitted from patient and public involvement; ethnographic methodologies, which allow for study of individuals who are not usually inclined to engage in research activities otherwise; the use of measures such as autovideography to allow participants to shape their own data freely; and mixed methods that allow for the inclusion of larger samples, such as Delphi studies used for questionnaire development.

### 1.5.2 Strengths and limitations

The findings in this review should be considered within the context of its strengths and limitations. To the authors’ knowledge, this is the first systematic review to examine users and caregivers’ understanding of recovery. The use of PRISMA guidelines and quality assessment of the studies added transparency and rigour to the research. However, research about recovery from the perspective of people of diverse backgrounds seemed to only start being documented in recent years. Despite applying a comprehensive search strategy, the evidence found in this review originated mainly from high-income, white-European populations due to a paucity of research in the field of recovery outside of these groups. Therefore, applicability of these findings outside of this context should be done with caution. Additionally, the proposed model of recovery could be strengthened in the future by researching grey literature or literature about concepts adjacent to recovery, such as studies which focused specifically on the notion of hope, empowerment or social inclusion.

## 1.6 Conclusion

The findings of this review provide context and depth to the construct of recovery, and add further evidence to emphasise the importance of social and clinical aspects of recovery. The comprehensive recovery model provided by this review offers a template for further research in the field and a guide for policy and practice development.

Evidence-based recovery research and practice relies on accurate representations of recovery goals and experiences in order to adequately address people’s needs. With sufficient attention to holistic models of recovery that represent the broad range of domains that interest users and carers, along with the promotion of their active participation, the recovery movement can continue towards fulfilling its commitment to have people with lived experience at the centre of decision-making in mental health.

## Supporting information

Supplementary File 1 Search strategy

Supplementary File 2 Inclusion exclusion criteria

Supplementary File 3 Risk of bias assessment

Supplementary File 4 Excluded articles

Supplementary File 5 Characteristics of included studies

## Data Availability

All data relevant to the study are included in the article or uploaded as supplementary information.

## Notes

### Competing Interest Statement

The authors have declared no competing interest.

### Funding Statement

This study was part of a PhD supported by National Institute for Health Research (NIHR) Collaboration for Leadership in Applied Health Research and Care South London (NIHR CLAHRC South London) at King’s College Hospital NHS Foundation Trust. The views expressed in this article are those of the author(s) and not necessarily those of the NHS, the NIHR, or the Department of Health and Social Care.

