## Supplementary File 1 Search strategy for "Recovery from severe mental health problems: A systematic review of service user and informal caregiver perspectives"

**Ovid (Embase, PsychINFO and Medline)**

1. (knowledge OR experienc* OR concept* OR narrative) mp.
2. AND ("mental disorder" OR "mental illness*" OR "mental problem*" OR "mental health" OR "mental disease*" OR "psych* disorder" OR "psych* illness*" OR "psych* problem*" OR "psych* health" OR "psych* disease*”)
3. AND (recover* OR wellbeing OR "personal recovery”)

LILACS and ScIELO

Terms used in Portuguese and Spanish to refer to recovery were added to the search to increase sensitivity to Latin American literature in these Latin American datasets.

1. (knowledge OR experienc* OR concept* OR narrative)
2. AND ("mental disorder" OR "mental illness*" OR "mental problem*" OR "mental health" OR "mental disease*" OR "psych* disorder" OR "psych* illness*" OR "psych* problem*" OR "psych* health" OR "psych* disease*”)
3. AND (recover* OR wellbeing OR "personal recovery” OR superaçao OR recuperaçao OR recuperación)

CINAHL

Due to obtaining no results with the strategy used in previous datasets, the search words were reduced to “recovery in mental health”
