## Supplementary File 2 Inclusion exclusion criteria for "Recovery from severe mental health problems: A systematic review of service user and informal caregiver perspectives"

Inclusion

- Focus of the research is recovery from mental disorders. Recovery understood as changes towards feeling well, reaching meaningful outcomes or experiencing a positive sense of self.
- As understood by service users and informal carers. Informal carer refers to people who provide unpaid care or support for people with mental disorders.
- Enquired through methodologies where participants’ perspectives were explored in an open-ended manor.
- No restrictions on the types of study design, publication date or location of the study.

Exclusion

Mental problems are not the primary condition:

- Participants have a known physical health condition or impediment.
- Participants are being studied because they were recently exposed to traumatic events.
- Participants’ primary problem is substance misuse.

The main outcome of the study is not knowledge, experiences or perceptions about personal recovery, but rather:

- Strategies to achieve a better recovery.
- Factors that mediate or moderate recovery.
- The experience of having a mental illness.
- Quality of care.
- Consequences of being an informal carer.
- Recovery assessment as an outcome of an intervention.
- Editorials / discussion / theoretical papers
- Dissertations and conference presentations.
