## Supplementary File 3 Risk of bias assessment for "Recovery from severe mental health problems: A systematic review of service user and informal caregiver perspectives"

Quality Assessment Checklist

Epistemological soundness

Are the research question/objectives clear?

Is a qualitative approach appropriate/justified to answer the research question?

Characteristics of the study

Are the sources of data /population adequately described?

Is sampling strategy described?

Is there coherence between objectives – results - discussion?

Are research questions and objectives clear and focused?

Has the relationship between researcher and participants been considered?
