## Supplementary File 4 Excluded articles for "Recovery from severe mental health problems: A systematic review of service user and informal caregiver perspectives"

##### Appendix 1 Articles excluded after full text revision. Citations and reasons for exclusion.

Studies excluded from the qualitative meta-narrative review in once full text articles had been accessed (n=109). Primary reasons for exclusion were studies focusing on the experience/aetiology of mental health problems, data not being presented separately for users and other populations, or the study limiting the definition of recovery by using measures with predetermined closed answers.

| **REFERENCE CITATION** | **REASON FOR EXCLUSION** |
| --- | --- |
| Acero González, Á. R., Cano-Prous, A., & Canga, A. (2016, August). Experiencias de la familia que convive con la enfermedad mental grave: un estudio cualitativo en Navarra. In Anales del Sistema Sanitario de Navarra (Vol. 39, No. 2, pp. 203-212). Gobierno de Navarra. Departamento de Salud. | Recovery defined as a process and what helps or hinders recovery. good for recovery. Recommendations for other families, family needs. |
| Adame, A. L., & Knudson, R. M. (2008). Recovery and the good life: How psychiatric survivors are revisioning the healing process. Journal of Humanistic Psychology, 48(2), 142-164. | Focus on commitment to political activism. . Focus only on one aspect of recovery. focus on commitment to political activism, social justice, |
| Agrest, M. (2016). First person narratives: Is there anything new in them for a mental health professional?. Vertex (Buenos Aires, Argentina), 27(128), 274-279. | Recovery defined without direct user or carer input. No user input. |
| Albrigtsen, V., Eskeland, B., & Mæhle, M. (2016). Ties of silence-Family lived experience of selective mutism in identical twins. Clinical child psychology and psychiatry, 21(2), 308-323. | Description of the experience of having mental health problems /illness. Focused on describing the treatment experience and explanation of the family and children of why the mental health problems occurred. |
| Anttila, K., Anttila, M., Kurki, M., Hätönen, H., Marttunen, M., & Välimäki, M. (2015). Concerns and hopes among adolescents attending adolescent psychiatric outpatient clinics. Child and Adolescent Mental Health, 20(2), 81-88. | Description of the experience and the aetiology of mental health problems /illness. Wrong population: "Adolescents with serious mental disorders such as psychotic depression (ICD-10 codes F32.3, F33.3), bipolar disorder (F31), substance abuse (F10–F19) or primary eating disorder (F50; ICD-10, 2013) were excluded, likewise those admitted to psychiatric hospital wards or involved in a brief intervention at an outpatient clinic (three appointments or less)." Focus on adolescent concerns, not what recovery means. |
| Aston, V., & Coffey, M. (2012). Recovery: what mental health nurses and service users say about the concept of recovery. *Journal of Psychiatric and Mental Health Nursing*, *19*(3), 257-263. | Results not presented separately for users, carers and other populations. Results not presented separately for service users and nurses. |
| Borge, L., & Fagermoen, M. S. (2008). Patients' core experiences of hospital treatment: Wholeness and self-worth in time and space. *Journal of Mental Health*, *17*(2), 193-205. | Recovery defined as a process and what helps or hinders recovery |
| Boumans, J., Baart, I., Widdershoven, G., & Kroon, H. (2017). Coping with psychotic-like experiences without receiving help from mental health care. A qualitative study. Psychosis, 9(1), 1-11. | Recovery defined as a process and what helps or hinders recovery. |
| Bradshaw, W., Armour, M. P., & Roseborough, D. (2007). Finding a place in the world: The experience of recovery from severe mental illness. *Qualitative Social Work*, *6*(1), 27-47. | Recovery defined as a process and what helps or hinders recovery. Barriers to recovery. |
| Çam, M. O., & Uğuryol, M. (2019). From Mental Disorder to Recovery: Cultural Effect. Current Approaches in Psychiatry/Psikiyatride Guncel Yaklasimlar, 11(5). | Fails to provide original literature and is more of a comprehensive review and oversight of the literature. |
| Brooks, G. W., & Deane, W. N. (1960). Attitudes of released chronic schizophrenic patients concerning illness and recovery as revealed by a structured post‐hospital interview. Journal of clinical psychology, 16(3), 259-264. | Recovery defined as a process and what helps or hinders recovery. what is good for recovery and when and how fast change happened. |
| Câmara, M. C., & Pereira, M. A. O. (2011). Percepções de transtorno mental de usuários da Estratégia Saúde da Família. Revista Gaúcha de Enfermagem, 730-737. | Description of the experience and/or the aetiology of mental health problems /illness. does not include concept of recovery, only disorder. |
| Carpenter-Song, E. A., Holcombe, B. D., Torrey, J., Hipolito, M. M. S., & Peterson, L. D. (2014). Recovery in a family context: Experiences of mothers with serious mental illnesses. Psychiatric rehabilitation journal, 37(3), 162. | Recovery defined as a process and what helps or hinders recovery. difficulties in the recovery experience of urban, low-income African American mothers. |
| Clarke, S., Oades, L. G., & Crowe, T. P. (2012). Recovery in mental health: A movement towards well-being and meaning in contrast to an avoidance of symptoms. Psychiatric Rehabilitation Journal, 35(4), 297. | Analysis limited to define recovery following a pre-established model of recovery or closed measure. only use of closed assessment measures |
| Clements, K. (2012). Participatory action research and photovoice in a psychiatric nursing/clubhouse collaboration exploring recovery narrative. Journal of Psychiatric and Mental Health Nursing, 19(9), 785-791. | Focus on instrument development and psychometric properties. Instrument development. description of the experience of using this method. |
| Cohen, O. (2005). How do we recover? An analysis of psychiatric survivor oral histories. *Journal of Humanistic Psychology*, *45*(3), 333-354. | Recovery defined as a process and what helps or hinders recovery. |
| Colquhoun, B., Lord, A., & Bacon, A. M. (2018). A Qualitative Evaluation of Recovery Processes Experienced by Mentally Disordered Offenders Following a Group Treatment Program. Journal of Forensic Psychology Research and Practice, 18(5), 352-373. | Main focus on dramatherapy and its impact on recovery, no mention of recovery as a separate topic. |
| Connell, J., Carlton, J., Grundy, A., Buck, E. T., Keetharuth, A. D., Ricketts, T., ... & Brazier, J. (2018). The importance of content and face validity in instrument development: lessons learnt from service users when developing the Recovering Quality of Life measure (ReQoL). Quality of Life Research, 27(7), 1893-1902. | Focus on Quality of Life for users and what life is like with a disease rather than recovery for them. What is a good question to ask in order to identify what Quality of Life means |
| D’Abundo, M., & Chally, P. (2004). Struggling with recovery: Participant perspectives on battling an eating disorder. *Qualitative Health Research*, *14*(8), 1094-1106. | Focus on eating disorders. Eating disorders have a different focus for recovery. |
| Davidson, L., Borg, M., Marin, I., Topor, A., Mezzina, R., & Sells, D. (2005). Processes of recovery in serious mental illness: Findings from a multinational study. *American Journal of Psychiatric Rehabilitation*, *8*(3), 177-201. | Recovery defined as a process and what helps or hinders recovery |
| de Almeida Colvero, L., Ide, C. A. C., & Rolim, M. A. (2004). Família e doença mental: a difícil convivência com a diferença. Revista da Escola de Enfermagem da USP, 38(2), 197-205. | Description of the experience and the aetiology of mental health problems /illness. Does not include concept of recovery, only disorder. |
| Dein, S. (2003). Psychogenic death: Individual effects of sorcery and taboo violation. *Mental Health, Religion & Culture*, *6*(3), 195-202. | Recovery defined without direct user input. No focus on recovery. User’s or informal carer’s views are not presented, only author interpretations of their story. |
| dos Santos Reinaldo, A. M., & Saeki, T. (2004). Ouvindo outras vozes: relato de familiares sobre o convívio com o paciente psiquiátrico. Revista da Escola de Enfermagem da USP, 38(4), 396-405. | Description of the experience and/or the aetiology of mental health problems /illness. no focus on recovery. Only focus on meaning of disorder and experience of having a disorder. |
| Eltaiba, N., & Harries, M. (2015). Reflections on recovery in mental health: Perspectives from a Muslim culture. *Social work in health care*, *54*(8), 725-737. | Recovery defined as a process and what helps or hinders recovery. Right question, wrong results. Recovery as a process. |
| Fernandez, M. E., Breen, L. J., & Simpson, T. A. (2014). Renegotiating identities: Experiences of loss and recovery for women with bipolar disorder. Qualitative health research, 24(7), 890-900. | Description of the experience and the aetiology of mental health problems /illness. no focus on recovery. Only focus on "loss" and the impact of being diagnosed with a disorder. |
| Furnham, A., & Kramers, M. (1989). Eating-problem patients' conceptions of normality. *The Journal of genetic psychology*, *150*(2), 147-153. | Focus on eating disorders. Recovery as symptom remission. Recovery different in eating disorders. |
| Goldsmith, L. P. (2012). A discursive approach to narrative accounts of hearing voices and recovery. Psychosis, 4(3), 235-245. | Description of the experience and the aetiology of mental health problems /illness. focus on the experience of disorder and explanations. |
| Gordon, S. E., Ellis, P. M., Siegert, R. J., & Walkey, F. H. (2014). Core dimensions of recovery: a psychometric analysis. Administration and Policy in Mental Health and Mental Health Services Research, 41(4), 535-542. | Analysis limited to define recovery following a pre-established model of recovery or closed measure quanti. Found through Voicing psychotic experiences: A reconsideration of recovery and diversity. Exclude because closed assesment measure. |
| Green, T., Batson, A., & Gudjonsson, G. (2011). The development and initial validation of a service-user led measure for recovery of mentally disordered offenders. Journal of Forensic Psychiatry & Psychology, 22(2), 252-265. | Focus on instrument development and psychometric properties. Recovery defined as a process and what helps or hinders recovery. no detail on development. Recovery as a process. |
| Gwinner, K., Knox, M., & Brough, M. (2013). Making sense of mental illness as a full human experience: Perspective of illness and recovery held by people with a mental illness living in the community. *Social Work in Mental Health*, *11*(2), 99-117.  Habhab, L (2016) Recovery experiences amongst Arab American Clubhouse Members: Examining the effects of Acculturation, Percieved Family Support, Stigma and Gender on mental Health recovery | Opinions about recovery as a concept (is recovery possible? Is it the right term?). Right research question, wrong results. Results talk about the acceptance of the word recovery.  Excluded for focussing on the effects of clubhouse on recovery – not on what recovery is to the user. |
| Henderson, A. R., & Cock, A. (2015). The responses of young people to their experiences of first-episode psychosis: Harnessing resilience. *Community mental health journal*, *51*(3), 322-328.  Hesamzadeh, A., Dalvandi, A., Maddah, S.B., Khoshknab, M.F., & Ahmadi,F. (2018). Family Caregivers’ experiences of Stroke Recovery Among Older Adults Living in Iran: A Qualitative Study. Iranian Red Crescent Medical Journal, 20(S1). | Description of the experience of having mental health problems /illness. The aim is to “build on existing knowledge and explore the responses of young people’s experience of a first episode of psychosis from the time their psychosis was detected until our contact with them”  Main focus on stroke recovery victims, recovery from mental disorders not written about |
| Hickey, J. E., Pryjmachuk, S., & Waterman, H. (2017). Exploring personal recovery in mental illness through an Arabic sociocultural lens. Journal of psychiatric and mental health nursing, 24(2-3), 163-170. | Overview definition of recovery and various definitions, no real definition as defined by users/carers themselves, more theoretical. |
| Higginson, S., & Mansell, W. (2008). What is the mechanism of psychological change? A qualitative analysis of six individuals who experienced personal change and recovery. *Psychology and Psychotherapy: Theory, Research and Practice*, *81*(3), 309-328. | Recovery defined as a process and what helps or hinders recovery. Wrong focus: explanation of how and why psychological change occurs. |
| Horsfall, D., Paton, J., & Carrington, A. (2017). Experiencing recovery: findings from a qualitative study into mental illness, self and place. Journal of Mental Health, 1-7.  Isobel, S. (2019). ‘In some ways it all helps but in some ways it doesn't’: The complexities of service users’ experiences of inpatient mental health care in Australia. *International journal of mental health nursing*, *28*(1), 105-116. | Recovery defined as a process and what helps or hinders recovery. "understand more about what is effective in the service system; what impacts living in a regional location might have on a person’s lived and service system experience and what people find helpful and supportive in their recovery journeys"  Main examination on how care during hospitalisation affects the participant, and the effects of admission to a clinical hospital |
| Iseselo, M. K., & Ambikile, J. S. (2020). Promoting Recovery in Mental Illness: The Perspectives of Patients, Caregivers, and Community Members in Dar es Salaam, Tanzania. Psychiatry Journal, 2020. | Primary focus on processes that help lead to recovery, lack of definition of actual recovery |
| Jacob, S., Munro, I., Taylor, B. J., & Griffiths, D. (2017). Mental health recovery: A review of the peer-reviewed published literature. Collegian, 24(1), 53-61. | Review of the literature with no original qualitative work being done however it does provide some interesting ideas with links to case studies. |
| Jacobson, N. (2003). Defining recovery: An interactionist analysis of mental health policy development, Wisconsin 1996-1999. *Qualitative Health Research*, *13*(3), 378-393. | Recovery defined without direct user or carer input. Wrong population: The members of the focus group are not described as users or informal carers. |
| Jensen, L. W., & Wadkins, T. A. (2007). Mental health success stories: Finding paths to recovery. *Issues in Mental Health Nursing*, *28*(4), 325-340. | Recovery defined as a process and what helps or hinders recovery. “The purpose of this study was to identify important factors, both formal and informal, that contributed to recovery from the perspective of persons with mental illnesses.” |
| Judge, A. M., Estroff, S. E., Perkins, D. O., & Penn, D. L. (2008). Recognizing and responding to early psychosis: a qualitative analysis of individual narratives. Psychiatric services, 59(1), 96-99. | Description of the experience of having mental health problems /illness Focus on illness recognition |
| Kelly, J., Gallagher, S., & McMahon, J. (2017). Developing a recovery college: a preliminary exercise in establishing regional readiness and community needs. Journal of Mental Health, 26(2), 150-155. | Results not presented separately for users, carers and other populations. mixed sample of users, community, staff and carers. |
| Keetharuth, A. D., Taylor Buck, E., Acquadro, C., Conway, K., Connell, J., Barkham, M., ... & Brazier, J. (2018). Integrating qualitative and quantitative data in the development of outcome measures: The case of the Recovering Quality of Life (ReQoL) measures in mental health populations. International journal of environmental research and public health, 15(7), 1342. | Focus on Quality of Life and how this affects users rather than recovery as a topic. |
| Kidd, S., Kenny, A., & McKinstry, C. (2015). The meaning of recovery in a regional mental health service: an action research study. Journal of Advanced Nursing, 71(1), 181-192. | Results not presented separately for users, carers and other populations. Results not presented separately for users, carers and staff. |
| Koenig, M (2017) Recovery in schizophrenia and sense of self. Annales Médico-psychologiques, revue psychiatrique, 175(8), 726-729 | No original research, an overview of previous ideas with some proposals of future work but no individual responses to what recovery means and definition |
| Kogstad, R. E., Ekeland, T. J., & Hummelvoll, J. K. (2011). In defence of a humanistic approach to mental health care: recovery processes investigated with the help of clients' narratives on turning points and processes of gradual change. Journal of psychiatric and mental health nursing, 18(6), 479-486. | Recovery defined as a process and what helps or hinders recovery. "The aim of this study is to improve the mental health nurses’ and other professionals’ understanding of different elements in change processes that support recovery, as experienced by mental health clients" |
| Kotake, R., Kanehara, A., Miyamoto, Y., Kumakura, Y., Sawada, U., Takano, A., ... & Kawakami, N. (2020). Reliability and validity of the Japanese version of the INSPIRE measure of staff support for personal recovery in community mental health service users in Japan. BMC psychiatry, 20(1), 51. | Main focus on INSPIRE and how it relates to the old ways and with no definition of recovery to the individual. |
| Lal, S., Ungar, M., Leggo, C., Malla, A., Frankish, J., & Suto, M. J. (2013). Well-being and engagement in valued activities: experiences of young people with psychosis. *OTJR: occupation, participation and health*, *33*(4), 190-197. | Recovery defined as a process and what helps or hinders recovery. Aim says, “well-being enhancing experiences”, but results are recovery as an outcome “activities perceived as being valuable to wellbeing”. |
| Larsen, T.: Lucy S.D.; (2018). Understanding the process of recovery from critical illness from the patient perspective: A constructivist grounded theory (Doctoral dissertation, The University of Western Ontario). | Theory paper with no actual recordings from individuals on recovery to them. Overview with theory generation and no set definition of recovery. |
| Lemos-Giráldez, S., García-Alvarez, L., Paino, M., Fonseca-Pedrero, E., Vallina-Fernández, O., Vallejo-Seco, G., ... & Barajas, A. (2015). Measuring stages of recovery from psychosis. Comprehensive psychiatry, 56, 51-58.  Leonhardt, B., Huling, K., Lysaker, P., Ratliff, K., Francis, M., & Breier, A. (2018, October). A Preliminary Exploration of the Relationship between Subjective and Objective Aspects of Recovery in a First Episode Psychosis Sample. In *EARLY INTERVENTION IN PSYCHIATRY* (Vol. 12, pp. 119-119). 111 RIVER ST, HOBOKEN 07030-5774, NJ USA: WILEY | Analysis limited to define recovery following a pre-established model of recovery or closed measure Recovery conceived only as the stages in the STORI. Recovery measures quantitatively.  Unable to find within an extensive search of literature |
| Liberman, R. P., Kopelowicz, A., Ventura, J., & Gutkind, D. (2002). Operational criteria and factors related to recovery from schizophrenia. *International Review of Psychiatry*, *14*(4), 256-272. | Results not presented separately for users, carers and other populations. Wrong population – The focus groups to validate a definition of recovery were formed by clinical researchers, mental health professionals and consumers. |
| Lloyd, R. (2007). Modeling community-based, self-help mental health rehabilitation reform. *Australasian Psychiatry*, *15*(1_suppl), S99-S103. | Results not presented separately for users, carers and other populations. Wrong population: the study included people with mental disorders, people with disabilities and people from the community. Results are not presented separately. |
| Lopes, T. S., Dahl, C. M., Serpa Jr, O. D. D., Leal, E. M., Campos, R. T. O., & Diaz, A. G. (2012). O processo de restabelecimento na perspectiva de pessoas com diagnóstico de transtornos do espectro esquizofrênico e de psiquiatras na rede pública de atenção psicossocial. Saúde e Sociedade, 21(3), 558-571. | Recovery defined as a process and what helps or hinders recovery. No focus on defining recovery. Focus on what users expect and what is good/bad for recovery. |
| Magalhães, C. D., de Mendonça Moscoso, J. T., de Souza Mitkiewicz, F., Wainstok, M. E. B., Fernandes, J. D. C. S., Marcos, G. L., & Tavares, M. C. (2013). " I am nuts, but networking": the qualification process for peer support work with mental health users in the psychosocial care network of Rio de Janeiro. Vertex (Buenos Aires, Argentina), 24(112), 445-454.  Mak, W. W., Chan, R.C., & Yau, S.S. (2018). Development and Validation of Attitudes towards Recovery Questionnaire across Chinese people in recovery, their family carers, ande service providers in Hong Kong. Psychiatry research, 267, 48-55. | Description of training in recovery for peer support workers. El objetivo de este artículo es describir el proceso de capacitación de usuarios de servicios de Salud Mental para trabajar como compañeros para la recuperación de otros usuarios de la red de atención psicosocial de la ciudad de Río de Janeiro.  Main focus on developing a new questionnaire, comparing it to other recovery questionnaires. No focus on what recovery means, more on the valdiity of scale. |
| Mansbach-Kleinfeld, I., Sasson, R., Shvarts, S., & Grinshpoon, A. (2007). What education means to people with psychiatric disabilities: A content analysis. *American Journal of Psychiatric Rehabilitation*, *10*(4), 301-316. | Focus only on one aspect of recovery. Recovery with a focus: understand what needs and values education fulfils for users. |
| Marin, I., Mezzina, R., Borg, M., Topor, A., STAECHELI LAWLESS, M. A. R. T. H. A., Sells, D., & Davidson, L. (2005). The person's role in recovery. *American Journal of Psychiatric Rehabilitation*, *8*(3), 223-242. | Recovery defined as a process and what helps or hinders recovery. |
| Marshall, S., Deane, F., Crowe, T., White, A., & Kavanagh, D. (2013). Carers’ hope, wellbeing and attitudes regarding recovery. *Community mental health journal*, *49*(3), 344-353. | Opinions about recovery as a concept (is recovery possible? Is it the right term?). “attitudes” only refers to “positive” or “negative” attitudes, not a definition of recovery. |
| Mead, S., & Copeland, M. E. (2000). What recovery means to us: Consumers' perspectives. *Community mental health journal*, *36*(3), 315-328. | Recovery defined as a process and what helps or hinders recovery. |
| Michalak, E. E., Yatham, L. N., Kolesar, S., & Lam, R. W. (2006). Bipolar disorder and quality of life: a patient-centered perspective. Quality of Life Research, 15(1), 25-37.  Minogue, J. L. (2015). *Subjective Experience of Recovery from Serious Mental Illness in Younger and Older Adults*(Doctoral dissertation, William James College) | Results not presented separately for users, carers and other populations. results not presented separately for users, carers and staff.  Focus on clubhouse members and how clubhouses help/hinder recovery, little focus on recovery to the individual |
| Moses, T. (2011). Parents’ conceptualization of adolescents’ mental health problems: Who adopts a psychiatric perspective and does it make a difference?. *Community mental health journal*, *47*(1), 67-81. | Description of the experience and/or the aetiology of mental health problems /illness. No focus on user recovery. |
| Muñoz González, L. A., Price Romero, Y. M., Reyes López, M., Ramírez, M., & Costa Stefanelli, M. (2010). Vivencia de los cuidadores familiares de adultos mayores que sufren depresión. Revista da Escola de Enfermagem da USP, 44(1). | Description of the experience and the aetiology of mental health problems /illness. no focus on recovery. Only focus on meaning of depression and the role of informal carers. |
| Myers, N. A., Smith, K., Pope, A., Alolayan, Y., Broussard, B., Haynes, N., & Compton, M. T. (2016). A mixed-methods study of the recovery concept,“a meaningful day,” in community mental health services for individuals with serious mental illnesses. *Community mental health journal*, *52*(7), 747-756. | Results not presented separately for users, carers and staff. |
| Nakamura, E., & Santos, J. Q. D. (2006). Depressão infantil: abordagem antropológica. Revista de saúde pública, 41, 53-60. | Description of the experience and/or the aetiology of mental health problems /illness. no focus on recovery. Only focus on meaning of depression. |
| Napo, F., Heinz, A., & Auckenthaler, A. (2012). Explanatory models and concepts of West African Malian patients with psychotic symptoms. *European Psychiatry*, *27*, S44-S49. | Description of the experience of having mental health problems /illness. No focus on user/carer definition of recovery. Focus on the experience of having mental health problems and theories of causes and facilitators. |
| Noiseux, S., & Ricard, N. (2008). Recovery as perceived by people with schizophrenia, family members and health professionals: A grounded theory. International Journal of Nursing Studies, 45(8), 1148-1162. | Results not presented separately for users, carers and other populations. |
| O’Keeffe, D., Hannigan, A., Doyle, R., Kinsella, A., Sheridan, A., Kelly, A., ... & Clarke, M. (2019). The iHOPE-20 study: relationships between and prospective predictors of remission, clinical recovery, personal recovery and resilience 20 years on from a first episode psychosis. Australian & New Zealand Journal of Psychiatry, 53(11), 1080-1092. | No description of recovery to the individual, explanation is explained but with no links to participants |
| Ospina, M., Zuvic, R., Ivanovic, F., & Lolas, S. (2009). Trastorno bipolar desde la perspectiva de los familiares. Trastor. ánimo, 5(1), 28-36. | Recovery defined as a process and what helps or hinders recovery and meaning of disorder diagnosis |
| Padgett, D. K., Smith, B. T., Choy-Brown, M., Tiderington, E., & Mercado, M. (2016). Trajectories of recovery among formerly homeless adults with serious mental illness. Psychiatric Services, 67(6), 610-614. | Analysis limited to define recovery following a pre-established model of recovery or closed measure focus on eight domains of recovery adapted from Whitley and Drake and what is good for recovery. |
| Palmquist, L., Patterson, S., O'Donovan, A., & Bradley, G. (2017). Protocol: A grounded theory of ‘recovery’—perspectives of adolescent users of mental health services. BMJ open, 7(7), e015161. | Focus on instrument development and psychometric properties. focus on instrument development. only protocol. Results not available. |
| Percy, M. L., Bullimore, P., & Baker, J. A. (2013). Voice hearer's perceptions of recovery: findings from a focus group at the Second World Hearing Voices Festival and Congress. *Journal of psychiatric and mental health nursing*, *20*(6), 564-568. | Results not presented separately for users, carers and other populations. Results not presented separately for users, carers and staff. |
| Pettersen, G., Thune‐Larsen, K. B., Wynn, R., & Rosenvinge, J. H. (2013). Eating disorders: challenges in the later phases of the recovery process. *Scandinavian journal of caring sciences*, *27*(1), 92-98. | Recovery defined as a process and what helps or hinders recovery. |
| Rashid, S. (2018). A Qualitative Study on the Process of Recovery and Resiliency in Trauma-based Combat Amputees (Doctoral dissertation, Alliant International University). | Unable to find anywhere for further research |
| Repper, J. (2000). Adjusting the focus of mental health nursing: Incorporating service users' experiences of recovery. *Journal of Mental Health*, *9*(6), 575-587. | Recovery defined as a process and what helps or hinders recovery. Summary of service user's views on recovery. |
| Romakkaniemi, M., & Kilpeläinen, A. (2015). The meaningful elements in recovering from major depression as a basis of developing social work in mental health services. Social Work in Mental Health, 13(5), 439-458. | Recovery defined as a process and what helps or hinders recovery. |
| Romano, D. M., McCay, E., Goering, P., Boydell, K., & Zipursky, R. (2010). Reshaping an enduring sense of self: the process of recovery from a first episode of schizophrenia. Early intervention in psychiatry, 4(3), 243-250. | Recovery defined as a process and what helps or hinders recovery process of recovery. |
| Romme, M., & Morris, M. (2013). The recovery process with hearing voices: accepting as well as exploring their emotional background through a supported process. *Psychosis*, *5*(3), 259-269. | Recovery defined as a process and what helps or hinders recovery. |
| Ryan, S., Rogers, A., & Lester, H. (2014). GPs’ and patients’ views on recovery from psychosis. *Mental Health Review Journal*, *19*(2), 99-109. | Results not presented separately for users, carers and other populations. Focus groups to define recovery done with GPs and patients together. Results not shown separately. |
| Saavedra Macías, F. J. (2011). Cómo encontrar un lugar en el mundo: explorando experiencias de recuperación de personas con trastornos mentales graves. História, Ciências, Saúde-Manguinhos, 18(1). | Historical review of the concept of recovery. |
| Saavedra, J. (2009). Schizophrenia, narrative and change: Andalusian care homes as novel sociocultural context. *Culture, Medicine, and Psychiatry*, *33*(2), 163-184. | Focus on effectiveness of treatment. |
| Salyers, M. P., Matthias, M. S., Sidenbender, S., & Green, A. (2013). Patient activation in schizophrenia: insights from stories of illness and recovery. *Administration and policy in mental health and mental health services research*, *40*(5), 419-427. | Focus on effectiveness of treatment. No focus on recovery, Patient activation refers to active participation in treatment. |
| Schon, U. K. (2009). How men and women in recovery give meaning to severe mental illness. *Journal of Mental Health*, *18*(5), 433-440. | Description of the experience of having mental health problems /illness meaning of illness |
| Schön, U. K. (2010). Recovery from severe mental illness, a gender perspective. Scandinavian Journal of Caring Sciences, 24(3), 557-564. | Recovery defined as a process and what helps or hinders recovery: "what the respondents regarded as having helped them to recover from severe mental illness" |
| Silva Pezo, M. C., Hoga Komura, L. A., & Stefanelli Costa, M. (2004). La depresión incluida en la historia de la familia. Texto & Contexto Enfermagem, 13(4). | Description of the experience and/or the aetiology of mental health problems /illness. focus on defining causes of depression from user and family perspective. |
| Singh, N. S., Jakhaia, N., Amonashvili, N., & Winch, P. J. (2016). “Finding a way out”: Case histories of mental health care-seeking and recovery among long-term internally displaced persons in Georgia. *Transcultural psychiatry*, *53*(2), 234-256. | Description of the experience of having mental health problems /illness Wrong focus- The paper seeks to understand how individuals living with mental health problems understand and manage their illness alongside the complexities of displacement. |
| Singla, N., Avasthi, A., & Grover, S. (2020). Recovery and its correlates in patients with schizophrenia. Asian Journal of Psychiatry, 102162. | Quantitative method used and lack of description of recovery and what it means to patients |
| Skärsäter, I., Dencker, K., Bergbom, I., Häggström, L., & Fridlund, B. (2003). Women's conceptions of coping with major depression in daily life: a qualitative, salutogenic approach. *Issues in Mental Health Nursing*, *24*(4), 419-439. | Recovery defined as a process and what helps or hinders recovery. |
| Song, L. Y., & Hsu, S. T. (2011). The development of the Stages of Recovery Scale for persons with persistent mental illness. *Research on Social Work Practice*, *21*(5), 572-581. | Recovery defined without direct user or carer input. no user input in the development of the scale. |
| Spaniol, L., Wewiorski, N. J., Gagne, C., & Anthony, W. A. (2002). The process of recovery from schizophrenia. *International Review of psychiatry*, *14*(4), 327-336. | Recovery defined as a process and what helps or hinders recovery. Focus on defining the experience of living with disability and what helps or hinders recovery. |
| Sravanti, L., & Kommu, J. V. S. (2020, January). Recovery Model of children and adolescents with obsessive-compulsive disorder: a qualitative study. In INDIAN JOURNAL OF PSYCHIATRY (Vol. 62, pp. S28-S28). WOLTERS KLUWER INDIA PVT LTD, A-202, 2ND FLR, QUBE, CTS NO 1498A-2 VILLAGE MAROL, ANDHERI EAST, MUMBAI, 400059, INDIA: WOLTERS KLUWER MEDKNOW PUBLICATIONS. | Unable to find paper for further examination |
| Stanton, J., & Simpson, A. I. (2006). The aftermath: Aspects of recovery described by perpetrators of maternal filicide committed in the context of severe mental illness. Behavioral sciences & the law, 24(1), 103-112. | Interviews with users in recovery that focus on another topic. No focus on mental illness recovery. |
| Suaalii-Sauni, T., Wheeler, A., Saafi, E., Robinson, G., Agnew, F., Warren, H., ... & Hingano, T. (2009). Exploration of Pacific perspectives of Pacific models of mental health service delivery in New Zealand. *Pacific Health Dialog*, *15*(1), 18-27. | Results not presented separately for users, carers and other populations. Focus on “pacific” services. |
| Swarbrick, M. A. (2013). Integrated care: wellness-oriented peer approaches: a key ingredient for integrated care. *Psychiatric Services*, *64*(8), 723-726. | Description of the experience of having mental health problems /illness Wrong focus: article not focused on the definition of wellness. |
| Thompson, E. H. (1989). Recovery networks and patient interpretations of mental illness. Journal of Community Psychology, 17(1), 5-17. | Description of the experience and/or the aetiology of mental health problems /illness. No focus on defining recovery. Focus on recovery networks and aetiology of mental disorders from user's view. |
| Torn, A. (2011). Chronotopes of madness and recovery: A challenge to narrative linearity. *Narrative Inquiry*, *21*(1), 130-150. | Analysis of time and space representation in the user ‘s narrative. No focus on the user recovery story. The main aim is to: examine the different ways in which time and space are represented in the narrative. |
| Tse, S., Murray, G., Chung, K. F., Davidson, L., Ng, K. L., & Yu, C. H. (2014). Exploring the recovery concept in bipolar disorder: a decision tree analysis of psychosocial correlates of recovery stages. *Bipolar disorders*, *16*(4), 366-377. | Analysis limited to define recovery following a pre-established model of recovery or closed measure Definition of recovery limited to the stages proposed in the Chinese-language Stages of Recovery Scale. |
| van Gestel-Timmermans, J. A. W. M., Brouwers, E. P. M., Bongers, I. L., van Assen, M. A. L. M., & van Nieuwenhuizen, C. (2012). Profiles of individually defined recovery of people with major psychiatric problems. International Journal of Social Psychiatry, 58(5), 521-531. | Recovery defined as a process and what helps or hinders recovery. |
| Waks, S., Scanlan, J. N., Berry, B., Schweizer, R., Hancock, N., & Honey, A. (2017). Outcomes identified and prioritised by consumers of Partners in Recovery: a consumer-led study. BMC psychiatry, 17(1), 338. | Focus on effectiveness of treatment. evaluation of an intervention |
| Watson, D. P., & Rollins, A. L. (2015). The meaning of recovery from co-occurring disorder: Views from consumers and staff members living and working in housing first programming. *International journal of mental health and addiction*, *13*(5), 635-649. | Focus on homelessness or addictions in co-occurring disorders. Unclear population. Focus on homelessness and addictions. |
| Whitwell, D. (1999). The myth of recovery from mental illness. Psychiatric Bulletin. | Opinions about recovery as a concept (is recovery possible? Is it the right term?). Discussion about whether recovery is possible. |
| Williams, C. C., & Collins, A. A. (1999). Defining new frameworks for psychosocial intervention. Psychiatry, 62(1), 61-78. | Description of the experience of having mental health problems /illness experience of illness, sense of self. |
| Wolstencroft, K., Oades, L., Caputi, P., & Andresen, R. (2010). Development of a structured interview schedule to assess stage of psychological recovery from enduring mental illness. *International Journal of Psychiatry in Clinical Practice*, *14*(3), 182-189. | Focus on instrument development and psychometric properties. No clear information about the recovery themes. Recovery as a process. Focus on instrument development. |
| Wood, S., Harrison, L. K., & Kucharska, J. (2017). Male professional footballers’ experiences of mental health difficulties and help-seeking. The Physician and sportsmedicine, 45(2), 120-128. | Description of the experience of having mental health problems /illness No focus on recovery. Focus on the experience of having mental health problems and barriers to help-seeking. |
| Yildiz, M., Erim, R., Soygur, H., Tural, U., Kiras, F., & Gules, E. (2018). Development and validation of the Subjective Recovery Assessment Scale for patients with schizophrenia. Psychiatry and Clinical Psychopharmacology, 28(2), 163-169. | Heavy focus on development of scale, no mention of recovery to the individual |
| Yip, K. S. (2003). Traditional Chinese religious beliefs and superstitions in delusions and hallucinations of Chinese schizophrenic patients. International Journal of Social Psychiatry, 49(2), 97-111. | Description of the experience of having mental health problems /illness no focus on recovery, focus on the experience of mental problems. |
| Zaraza-Morales, D. R., & Hernández-Holguín, D. M. (2017). Encerrado a oscuras: significado de vivir con esquizofrenia para diagnosticados y sus cuidadores, Medellín-Colombia. Aquichan, 17(3). | Description of the experience and the aetiology of mental health problems /illness. no focus on recovery. focus on the experience of having mental problems. |
