## Supplementary File 5 Characteristics of included studies for "Recovery from severe mental health problems: A systematic review of service user and informal caregiver perspectives"

##### Supplementary File 5. Citation and characteristics of included studies

This appendix lists the references for all papers (n=62) included in the meta-narrative review.

| Study | Recovery paradigm | Setting (Country, City, Site) | Population studied | Data collection method | PPI | User outcome recovery themes | Carer outcome recovery themes | Study Quality Score* |
| --- | --- | --- | --- | --- | --- | --- | --- | --- |
| Adame & Knudson (2007) | Political recovery | USA, Western Massachusetts and Eastern Ohio, psychiatric survivor community | User N=4 | interview |  | empowerment, recovery as connection, peace of mind, control, autonomy, empowerment, dropping de label |  | 3 |
| Armour et al. (2009) | Social recovery | USA, Minnesota, non-profit community mental health program | User N=9 (5 females 4 males)  Age 25-54  African American  Severe mental illness  Four months service use | semi-structured interviews |  | being normal, recovery in the eyes of others, economic stability, connection. Woman users: being allowed to take risks |  | 4 |
| Basso et al. (2016) | USA consumer/ survivor movement  Social recovery  Political recovery | Italy, community mental health services throughout | Users N=82 Carers N=45  Age 24-65  Psychosis  Education 3-25 years  In symptomatic remission, good ability to sustain social role | interviews, focus group | PAR | personal growth, being happy, control, autonomy, embracing the label, participation | Clinical recovery* limited information | 4 |
| Bird et al. (2014) | REFOCUS-CHIME | UK, England, London, NHS trust | User N=48 (28 male, 20 female)  Age 18-65  White British, White Irish, white other, black - African, black - Caribbean, black other, Asian - Indian, Asian - Pakistani, Asian other, mixed race.  Sever mental illness | focus groups |  | connection, new identity, old identity, participation, empowerment |  | 4.5 |
| Borg & Davidson (2008) | Social recovery | x | User N=13 (7 women 6 men) Age 26-54 Psychosis, Manic depression, paranoia Higher education Employment 2 ordinary jobs, 11 disability payments and working part-time 3 recovered 2 heard voices | interview | PAR | being normal, everyday life activities, having economic stability, living in the present, control |  | 6 |
| Brijnath (2015) | REFOCUS-CHIME  USA consumer/ survivor movement | Australia, Melbourne | User N=58 (35 female 23 male)  Age 40.9 (SD 15.9)  Anglo-Saxon Celtic Australian, Indian immigrants  Depression  Full time Australians 23.3%, Indians 39,3% | interview |  | participation, recovery in everyday life activities, embracing the label-medication, dropping the label-medication, personal growth, empowerment, control, economic stability. Anglo participants: knowledge about mental health, embracing the label |  | 6 |
| Brooke-Sumner et al. (2014) | USA consumer/ survivor movement | South Africa, primary healthcare clinic and NGO | Users + carers N=9, 5 male 4 female  Age 21-59  Schizophrenia  2 no schooling, 3 primary education, 4 post-secondary  1 employed, 7 unemployed  Symptoms well managed | semi-structured individual qualitative interviews |  | x | economic stability, participation - paid or unpaid productive activity | 5.5 |
| Castillo et al. (2013) | REFOCUS-CHIME  USA consumer/ survivor movement | UK, Essex | User N=66  Personality disorder | focus groups and individual interviews | PAR | personal growth, recovery in everyday activities, being sensitive to other's needs, participation - paid or unpaid productive activity, being happy, education |  | 5 |
| Daley et al. (2013) | REFOCUS-CHIME | UK, South London, older people’s mental health service | User N=38  Over 65 years of age  Severe mental illness + dementia | interviews |  | old identity, personal growth, control. Dementia: recovery in everyday life activities |  | 6 |
| Davidson et al. (2010) | Social recovery | USA, Connecticut, local mental health clinics and psychosocial clubs | User N=30  Unemployed | interview | PAR | participation, old identity, formal education, autonomy, economic stability, being happy |  | 6 |
| Davis (1967) | Social recovery | USA, short treatment ward | User N~18 all women  Age 17-60  Cross-diagnostic  Acute symptoms, on drug therapy | Interviews, field notes. Observations of participants over 3 months. |  | good appearance, recovery in the eyes of others, old identity, being sensitive to other's needs, living in the present, co-construction |  | 5 |
| de Jager et al. (2016) | REFOCUS-CHIME | Australia, Hearing Voices Network NSW (HVNNSW) and Australian Schizophrenia Research Bank (ASRB) | User N=11 7 women 4 men  Age 23-63  White and 1 Asian  Psychosis  5 paid employment, 3 retired, 2 disability pension  Advanced recovery stage | Narrative Inquiry. semi structured interviews | user participants validated results | new identity, control, being normal, old identity, participation, connection, personal growth, knowledge about mental health |  | 4 |
| Engqvist & Nilsson (2014) | USA consumer/ survivor movement | Sweden | User N=13 female  Carer N=6, 4 female 2 male  Age 44-65  Post-partum psychosis  Recovered | interviews |  | control, being happy, connection, being sensitive to other's needs, everyday life activities - sleep | x | 5 |
| Fullagar & O'Brien (2014) | Political recovery | Australia, metropolitan and rural Queensland, and metropolitan and rural New South Wales | User N=31 female  Age 35-49  Anglo-Celtic background, middle European origins  Depression  Self-identified as recovered, 21 still on medication | Interview |  | embracing the label, dropping the label, being allowed to take risks |  | 5 |
| Gillard et al. (2015) | REFOCUS-CHIME  USA consumer/ survivor movement | UK, trust | User N=6 3 men 3 women  Age 26-65  White or other  Personality disorder  Attending peer support group 2+years | interview | service user researchers as part of the team | participation, personal growth, connection, formal education, control, being happy |  | 5 |
| Gopal et al (2020) | Consumer/survivor movement | Outpatient department Chennai, India, Schizophrenia Research Foundation | 100 clients with schizophrenia and 80 caregivers, 55% were males. the subjects were employed (32%) and unemployed (31%). (74%) were from urban background; 53.8% of the caregivers were males and 47% were parents and 24% were spouses. The subjects had a mean duration of illness as 11.4 years. Among the subjects, 43% of them were in remission | Snowballing technique, interview | Mental heath experts asks to comment on usage of words/phrases | Users: absence of symptoms, independence | Caregivers: free from relapses, expressing emotions, understood as elimination of symptoms | 4.5 |
| Güner (2014) | Social recovery  Bio-medical recovery | Turkey, Istanbul, schizophrenia patient associations | User N=9, 8 male 1 female  Age 27-54  Schizophrenia  4 university graduates  Welfare assistance | In-depth interviews |  | Clinical recovery, connection - getting married, economic stability, participation |  | 5 |
| Guzman et al. (2019) | Personal recovery based on defense of civil rights of people USA consumer/survivor movement | Talcahuano Concepcion in Chile | 16 participants; 7 females, bipolar/schizophrenia | Semi-structured interviews | Interviews coded by pairs with one research present during interview and another who was not. Review of coding | Personal development, social inclusion of participant (carrying out activities, absence of symptoms). Biomedical perspective |  | 5 |
| Hancock et al (2018) | REFOCUS-CHIME, recovery is seen as an individualised, ongoing journey towards living a life of personal meaning | Sydney, Australia | 13 consumers (11 female, 2 male), all had a severe and persistent mental illness with a complicating co-morbidity, including physical ill-health or addiction. | Semi-structured interview | Third author independently reviewed all coding | Engaging challenge of recovery, grieving for what was and what could have been, seeking and finding hope, connecting with support |  | 4 |
| Jacob et al. (2015) | REFOCUS-CHIME  USA consumer/ survivor movement | Australia, Victoria, Area Mental Health Service in | Users N=9, 6 male 3 female  Carer N=8, 2 male 6 female  User age 48, carer 63  Schizophrenia, bipolar  Currently in acute inpatient units | qualitative interviews |  | personal growth, embracing the label, control, being happy, being normal, peace of mind, autonomy, clinical recovery, participation | recovery is impossible, clinical recovery, recovery of old identity, autonomy | 5 |
| Kartalova-O'Doherty & Tedstone Doherty (2010) | USA consumer/ survivor movement | Ireland, two urban and rural counties, Peer support groups and MHS | User N=15, 9 male 6 female  Age 47.1  Schizophrenia, mood disorder, anxiety | Individual open-ended interviews | user participants validated results | peace of mind, personal growth, connection, control, being sensitive to other's needs, participation, being happy, economic stability, formal education |  | 5 |
| Katsakou et al. (2012) | REFOCUS-CHIME  USA consumer/ survivor movement | UK, East London, secondary mental health services | User N=48, 39 female 9 male  Age 36.5 (SD 10)  White, black and Asian  Bipolar disorder and comorbid diagnoses  unemployed 77%, voluntary work 6%, employed 17%  63% most life goals achieved | interview | service user researchers as part of the team | personal growth, control, connection, economic stability, being normal, clinical recovery |  | 6 |
| Kidd et al. (2014) | USA consumer/ survivor movement | Canada, Toronto, MHS | User N=6 female Age 31-53 Black African, Black African Canadian, Black Caribbean and South Asian Severe mental illness Average 12 years of education | in-depth arts-based qualitative mediums | user participants validated results | empowerment, being sensitive to other's needs, participation, being allowed to take risks, embracing the label |  | 6 |
| Kverme et al. (2019) | USA Consumer/Survivor movement | Norway | 12 participants, all female, 21-36, Bipolar Disorder | Individual in depth interviews | Review of results, critical auditor | Moving towards connectedness, honesty, daring to belong, making room for recovery, learning to hold ones own |  | 7 |
| Lal et al. (2014) | REFOCUS-CHIME | Canada, West coast urban, early intervention program for psychoses | User N=17 71% male Young, mean 22 years European, other (e.g. First Nations, Asian) Psychosis most unemployed and not completed high school | semi-structured interviews and participant-photography elicited focus groups |  | control, connection, recovery of old identity, new identity, being normal |  | 6 |
| Landeweer et al. (2009) | Bio-medical recovery | Netherlands, Amsterdam and Maastricht, clinical psychiatry facilities. | User N=1 female | Open interview, case analysis |  | recovery in the eyes of others, connection, autonomy |  | 6 |
| Law & Morrison (2014) | REFOCUS-CHIME  USA consumer/ survivor movement | UK, North West of England, 7 NHS mental health trusts in the | User N=381  50% female Most common age range 40-49 Psychosis | Delphi method | user advisory committee | economic stability, personal growth, being happy, overcoming barriers, empowerment, knowledge about mental health |  | 6 |
| Law et al. (2020) | REFOCUS-CHIME, recovery as a journey, self, social, family. Self-belief, responsibility, hope | Two different NHS Trusts across the UK | 23 participants, 18 females, 14-25yrs old, 20 identified as white | Semi-structured interviews | Inter-rater reliability | Independence, not relying on others, reduction of symptoms, individual ongoing journey towards stability |  | 7 |
| Leavey (2009) | Social recovery | Canada, urban community, psychosocial rehabilitation centre for youth | User N=13, 7 male 6 female Young, 17-23 years schizophrenia, psychosis, delusional disorder, obsessive-compulsive disorder, depression, suicidal ideation, eating disorder, anxiety disorder, bipolar disorder, mood disorder, post-traumatic stress disorder, personality disorders, and learning disabilities.  Taking medication | Semi-structured interview | user participants validated results | embracing the label, new identity, personal growth |  | 6 |
| Lee et al (2020) | SAHMSA | MidWestern US State | 19 individuals, 47.4% were male, mean age 46.68, 17 identified as White | Semi-structured interviews | Discussed amongst team to review established codes | Having a different relationship with symptoms, accepting neg. emotions, learning ways to cope, ownership of issues, hope, positivity. |  | 5 |
| Mananita et al. (2011) | USA consumer/ survivor movement | USA, Washington, DC, recovery communities. | User 75% female 47.11 average age | Focus groups |  | recovery s embracing the label, living in the present, everyday life activities, personal growth |  | 7 |
| McCabe et al. (2018) | Bio-Medical | North West England | White british Schizophrenic 9 Males  20-42yrs old | Interview | Supervision by sunior researcher who encouraged further reflection | Living in the community, importance of professionals, not a single state but a complex mix, attributed it to what the staff thought. |  | 6 |
| Mccauley et al. (2017) | REFOCUS-CHIME  USA consumer/ survivor movement | Northern Ireland, deprived cities | User Young adults | Discussion groups | user advisory committee | self-discovery, development of coping mechanisms, turning painful experiences into positive, finding purpose |  | 7 |
| Mezey et al. (2010) | USA consumer/ survivor movement | UK, England, London, medium secure unit | User N=10, 8 men 2 women  Age 24-56  White, clack and minority Schizophrenia detained in secure psychiatric facilities for 1-11 years | interview | service user researcher as part of the team and user advisory committee | personal growth, participation, recovery in the eyes of others, economic stability, connection, control, being normal |  | 7 |
| Milbourn et al. (2014) | REFOCUS-CHIME | Western Australia | User N=11, 8 men, 3 women Age 27-53 Psychosis Unemployed, receiving disability pension Hard to engage | Interview and field notes. longitudinal 12 months |  | being happy, looking into the future, economic stability, recovery in the eyes of others, control |  | 7 |
| Mizock et al. (2014) | USA consumer/ survivor movement | USA, Massachusetts northeast psychosocial rehab centre. | User N=23 | photovoice |  | personal growth, co-construction of recovery, being happy, looking into the future, control, connection, autonomy |  | 6.5 |
| Mizuno et al. (2015) | USA consumer/ survivor movement | Japan, psychiatric facilities | User N=16, 11 male, 5 female Age 43.8 (SD 10.5) Schizophrenia  Employment experience: yes 56%, none 43%  Duration of illness 22.4 years (9.1SD) | interview |  | participation, autonomy, knowledge about mental health, economic stability, clinical recovery |  | 6 |
| Moltu et al. (2017) | REFOCUS-CHIME  USA consumer/ survivor movement | Norway, Førde (Western region), public hospital | User N=50  heterogeneous trans-diagnostic sample | focus groups and individual interviews | user researchers as part of the team | control, recovery in the eyes of others, co-constructed |  | 7 |
| Moxham et al. (2017) | USA consumer/ survivor movement  Social recovery | Australia, New South Wales, Recovery camp | User N=27, 17 female 10 male  Age 22-63 Serious mental illness: Depression, bipolar, schizophrenia, anxiety, schizo-affective disorder, alcohol addiction, PTSD and borderline personality Stable living in the community | all the consumers were invited to write down up to five goals that they wished to achieve during the week. |  | connection, control, overcoming barriers |  | 7 |
| Nowak et al. (2017) | CHIME, bio-medical. | Warsaw, Poland | 28 participants, mean age was 43.78 year, majority were men (56.60 %), single (67.90 %), with vocational education (53.60 %), and unemployed—receiving disability benefits or pension (74.10 %). 28.60 % had a university degree. | Focus Group Discussion | Inter-rater reliability | Psychological recovery, clinical understanding of recovery, improving relationships with others, wellness strategies, support systems |  | 7 |
| Nxumalo Ngubane et al (2019) | Consumer/Survivor taking control of ones life | Swaziland | User N=15  21-70yrs old  Female | Face to face interviews were conducted |  | Having faith in self,medicine,religion, being useful, families and sig. others play an important role |  | 7 |
| Ochocka et al. (2005) | USA consumer/ survivor movement | Canada, Ontario, Consumer/survivor initiatives in community mental health | User N=28 | in-depth interviews. Baseline, 9 month and 18month | PAR | personal growth, connection, overcoming barriers, empowerment, control, autonomy, being happy |  | 4 |
| Petros et al. (2016) | REFOCUS-CHIME | USA, Philadelphia, recovery education program | User N=6  Age 18+  Severe mental illness Graduates of recovery education class | Autovideography |  | connection, recover as empowerment, participation, recovery a looking to the future, recovery of old identity, recovery of new identity |  | 4.5 |
| Piat et al (2009) | USA consumer/ survivor movement | Canada, Montreal MHS, Quebec community housing organisation, and Ontario mental health association. | User N=60, 52% female Mean age 43.6 (SD 8.96) Severe mental illness 50% high school graduates 45% current paid or unpaid work, 93% worked during lifetime 95% on medication, 72% not hospitalised in previous year | Interview | User advisory committee | clinical recovery, recovery of old identity, control, personal growth |  | 5.5 |
| Pitt et al. (2007) | USA consumer/ survivor movement | Britain, mental health groups | User N=7, 5 male 2 female  Age 18-65 White and 1 mixed race Psychosis | semi-structured interviews | user-led research | personal growth, knowledge about mental health, empowerment, autonomy, control, dropping the label, participation, connection, risks, happy, other's needs, co-construction |  | 5.5 |
| Ridge & Ziebland (2006) | No recovery paradigm | UK, diverse locations and settings | User N=38, 22 women 16 men  Age 18-66  White and of British ethnicity; Black, Asian, Southern European, Northern European, and American depression | video- and audiorecorded interviews |  | clinical recovery, personal growth, embracing the label, new identity |  | 5.5 |
| Ridgeway  (2001) | USA consumer/ survivor movement  Political recovery | USA | User N=4 | recovery narratives |  | looking to the future, recovery in the eyes of others, participation, control, dropping the label, personal growth, connection |  | 5.5 |
| Santos et al (2018) | REFOCUS-CHME | Southern California, usa, outpatient centre | 120 people 60 consumers - 40 male, mean age: 39.4; 60 caregivers - 11 male,mean age 55.12 Mexican origin willing to participate in the study. | Assessments followed by semi-structured interviews at baseline and post baseline 9 months later |  | , independence, self responsibility, empowerment, effect change in their life | Social relationships | 5 |
| Sheperd et al. (2017) | REFOCUS-CHIME | UK, North England, community hospitals, community mental health team bases, local prisons | User N=41 White or not comment  Personality disorder  Most not currently working | qualitative interviews |  | personal growth, control, embracing the label, looking to the future, dropping the label |  | 6.5 |
| Simonds et al. (2014) | Social recovery | UK, England, CAMHS | User + carer N=21, 7 girls 2 boys and 12 mothers Age users 14-16, carers 35-54 White British obsessive–compulsive disorder, panic disorder and generalized anxiety disorder and depression Carers professional roles, one unemployed Users 8 still in CAMHS, 4 discharged | interview | user advisory committee | being normal, new identity, personal growth, formal education, connection, economic stability, being happy, | being happy, clinical recovery, control, autonomy | 6.5 |
| Svanberg et al. (2010) | USA consumer/ survivor movement | West of Scotland, social firms | User N=16, 15 men Age 19-64 bipolar, depression, psychosis, comorbid with anxiety and addictions | interview | user participants validated results | looking to the future, control |  | 5 |
| Thornhill et al. (2004) | USA consumer/ survivor movement  Social recovery | UK, unspecified | User N=15, 9 women Age 30-70 White European and Asian Psychosis recovered | interview | user researchers as part of the team | empowerment, control, personal growth, knowledge about mental health, embracing the label |  | 4.5 |
| Todd et al. (2012) | USA consumer/ survivor movement | UK, North West England | User N=12, 7 male  Average age 42  White British  Bipolar  Average days since last episode 280 | Focus groups |  | being normal, new identity, control, risks, autonomy, overcoming barriers |  | 5.5 |
| Tofthagen et al. (2017) | Perkins & Slade distinguish from recovery from illness and recovering in life USA consumer/survivor movement | Norway | 8 N=8, (7 female), mean age of 36 | Eight transcribed interviews |  | Prolonged learning process, choose other actions, to attend to ones basic physical needs, verbally express inner pain |  | 5 |
| Tweedell et al. (2004) | USA consumer/ survivor movement | Canada, Ontario, psychiatric facility | Carer N=18  9 families: 5 parents, 1 mother, 1 spouse, 4 sibling in laws, 3 sisters, 4 brothers Age 27-78 Caucasian Schizophrenia 3-15 years minimal relief from symptoms | interview, 3 in one year |  |  | Being normal, personal growth, control, autonomy | 5.5 |
| Vander Kooji (2009) | Social recovery | x | Standardised dietary questions from User N=3, 2 female  Schizophrenia, depression, bipolar, anxiety Stable enough to participate | songs written by adult music therapy and Open-ended interviews |  | risks, peace of mind, happy, future |  | 5.5 |
| Whitley et al. (2016) | REFOCUS-CHIME | Canada, Montreal, psychiatric outpatient clinics. | User N=47, 24 mean Euro-Canadian, Caribbean-Canadian Severe mental illness | semi-structured interview |  | everyday activities, connection, participation, future |  | 5.5 |
| Wood et al. (2010) | Social recovery | UK, Manchester, NHS | User N=8, 6 male  Age 24-35  Psychosis Delusions or hallucinations in last 12 months | q-methodology, interviews | service user researcher as part of the team | clinical recovery, overcoming barriers, personal growth, control, economic stability, connection |  | 6.5 |
| Wood et al. (2013) | other-mixed | UK, Manchester, NHS | User N=40, 25 male Age 18-65 Mainly white Psychosis 18 Primary school education, 22 further education  Unemployed 35, employed 2 | semi-structured interview | service user researcher as part of the team | co-construction, old identity - occupational, personal growth, future |  | 6.5 |
| Yarborough et al. (2016) | REFOCUS-CHIME  USA consumer/ survivor movement | USA, Oregon and Washington state, integrated health plan (private) | User N=177 52% female  94%white  Schizophrenia, bipolar, affective psychosis | semi structured interviews baseline 12- and 24-months | user researchers as part of the team | economic stability, recovery in everyday activities, knowledge about mental health, control, overcoming barriers, being happy |  | 3.5 |
| Young & Ensing (1999) | USA consumer/ survivor movement | x | User N=18, 12 female  Age 26-59  African American, European-American bipolar, schizophrenia, anxiety, major depression, schizo-affective, psychotic depression, borderline personality, PTSD, Claustrophobia, bulimarexia, mental retardation. | interviews and focus groups |  | overcoming barriers, embracing label, old identity, empowerment, personal growth, connection, control, being normal, peace of mind, economic stability, clinical recovery |  | 5.5 |
| Yuen et al. (2019) | Leamy et al., involvement of families | Hong Kong | Carers N=14  11 female, mean age 54  Chinese | Semi-structured interviews with open ended questions |  |  | Recovery = ‘living a normal life’. Maintain long emotionally stable periods, reconnecting with society, observable signs-be independent, going out | 6 |
